# The impact of the SARS-CoV-2 pandemic on healthcare provision in Italy to non-COVID patients: a systematic review

**DOI:** 10.1101/2020.09.17.20192088

**Authors:** Lugli Gianmarco, Ottaviani Matteo Maria, Botta Annarita, Ascione Guido, Bruschi Alessandro, Cagnazzo Federico, Zammarchi Lorenzo, Romagnani Paola, Portaluri Tommaso

## Abstract

**Background:** Italy has been one of the countries most affected by the SARS-CoV-2 pandemic and the regional healthcare system has had to quickly adapt its organization to meet the needs of infected patients. This has led to a drastic change in the routine management of non-communicable diseases with a potential long-term impact on patient health care. We investigated the management of non-COVID-19 patients across all medical specialties during the pandemic in Italy.

**Methods:** A PRISMA guideline-based systematic review of the available literature was performed using PubMed, Embase, and Scopus, restricting the search to the main outbreak period in Italy (from 20 February to 22 June, 2020). We selected articles in English or Italian that detailed changes in the Italian hospital care for non-COVID-19 patients due to the pandemic. Our keywords included all medical specialties in combination with our geographical focus (Italy) and COVID-19.

**Findings:** Of the 4643 potentially eligible studies identified by the search, 247 studies were included in the systematic review. A decrease in the management of emergencies in non-COVID patients was found together with an increase in mortality. Similarly, non-deferrable conditions met a tendency toward decreased diagnosis. All specialties have been affected by the reorganization of healthcare provision in the hub-and-spoke system and have benefited from telemedicine during the pandemic.

**Interpretation:** Our work highlights the changes taking place in the Italian public healthcare system in order to tackle the developing health crisis due to the COVID-19 pandemic. The findings of our review may be useful to analyze future directions for the healthcare system in the case of new pandemic scenarios.

## Introduction

Since the first case of the novel coronavirus (COVID-19) was reported in Wuhan, China in December 2019, viral infection spread at an alarming rate worldwide. On January 30, 2020, the World Health Organization (WHO) described COVID-19 as a Public Health Emergency of International Concern, and by March 11, 2020, it was officially declared a pandemic.^1^ Italy was the first European country to be affected by COVID-19 with the first case being diagnosed on 20 February in a man living in the province of Lodi (NorthWest Italy).^2^ The epidemic went on to affect all regions in Italy, with higher incidence rates in the north. The peak of the COVID-19 epidemic in Italy was reached in the last week of March with over 5500 new cases per day.

There has been a gradual decline since then as a result of strict containment measures that shaped the Italian lockdown phase. However, especially during the first phase of the epidemic, the outbreak put the Italian National Health System (Servizio Sanitario Nazionale, SSN) under unprecedented pressure.

In an attempt to direct the available resources at counteracting and limiting the effects of the pandemic, deferrable and non-urgent medical activities were suspended. On the other hand, patients with life-threatening conditions, such as myocardial infarction and stroke, or chronic conditions, such as diabetes, retained the right to their medical needs being met.

In these circumstances, several medical domains have been constrained by different resource allocations with unpredictable long-term consequences on patient health care.^3,4,5^

Here, we present a systematic review of the literature which illustrates the direct and indirect effects of the COVID-19 pandemic on the management of non-COVID patients across all medical specialties.

## Methods

This systematic review was performed in accordance with PRISMA guidelines.^6^ The search was conducted on 22 June 2020 on three different databases: PubMed, Embase, and Scopus without any date restriction. All the keywords were investigated within the title and abstract in both “AND” and “OR” combinations. Our keywords included all medical specialties (and potential synonyms) in combination with our geographical focus (Italy or Italian) and COVID-19. The full search strategy is reported in Table 1, Supplementary Material.

**Table 1.**
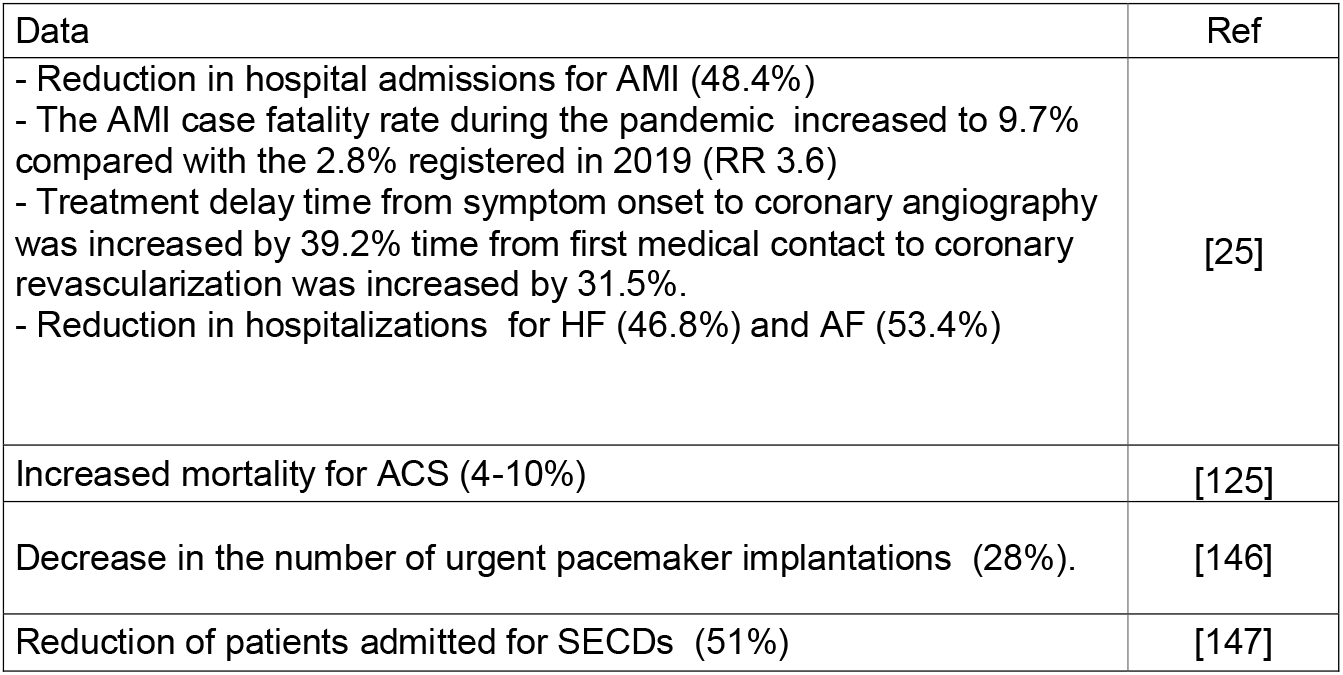
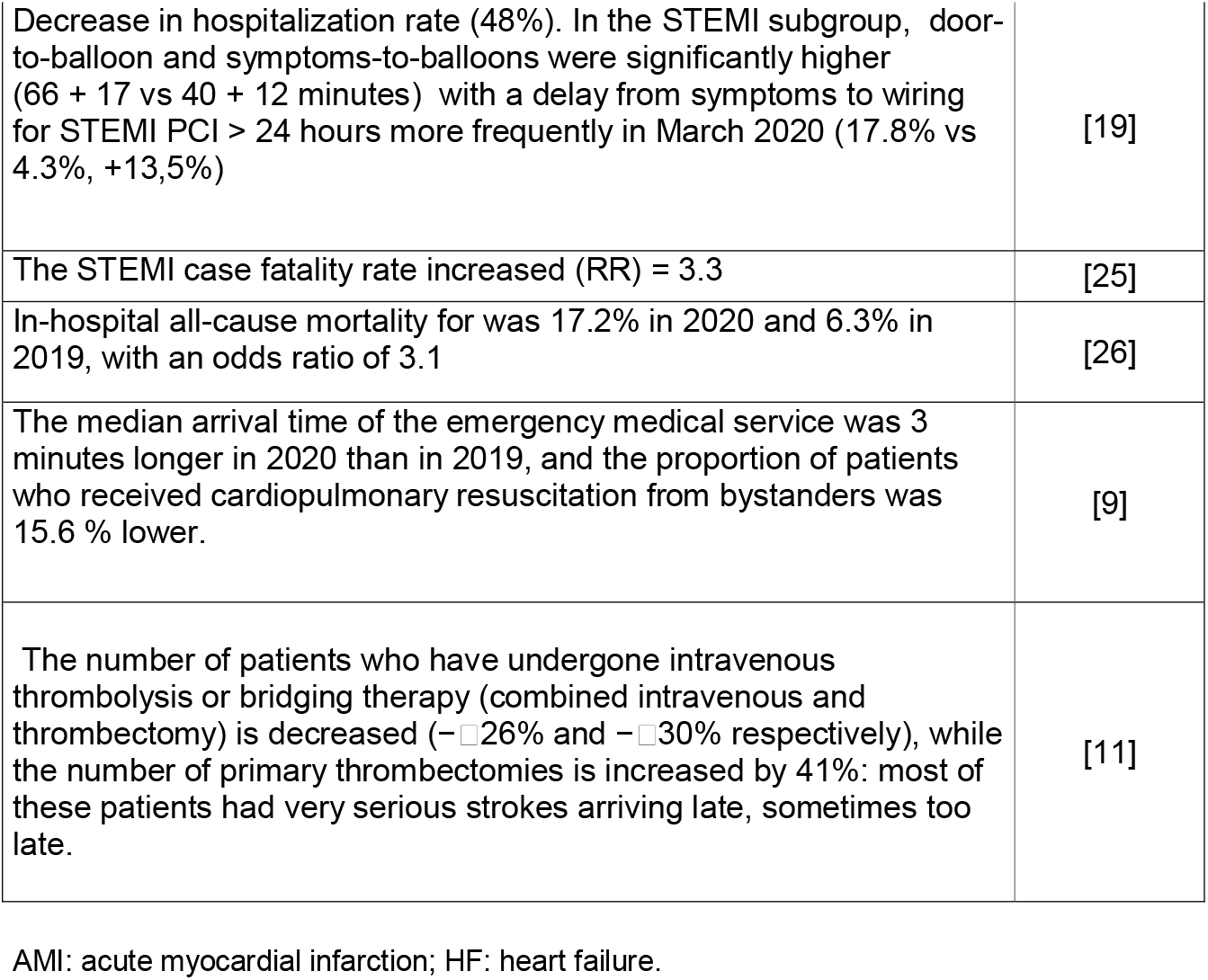
Quantitative data in cardiovascular emergencies.

### Selection of the studies

The literature search returned original papers published between 1979 and 2020 – especially for the keyword “coronavirus”. Since our focus was the novel severe acute respiratory syndrome coronavirus 2 (SARS-CoV-2) and the first positive case in Italy was detected on 20 February, the literature search was restricted to the period from 20 February to 25 June, 2020. The databases were queried via an R script on their respective APIs, checked and cleaned for duplicates (via title, DOI and/or database id), and exported into Excel.

In the second stage, studies were selected based on their titles and abstracts: each study was independently evaluated by three different raters (AnB, GA, GL). When there was a lack of agreement among the screeners, ensemble majority voting was used for the final decision. The full texts of the selected papers were thus analyzed by two reviewers in terms of relevance and inclusion/exclusion criteria (MMO and GA). When these reviewers disagreed over the inclusion or exclusion of a paper, a third reviewer was responsible for the final decision (GL). In addition, the reference lists of selected papers were reviewed in order to find pertinent studies not identified during the initial search.

### Inclusion and exclusion criteria

The simultaneous co-occurrence of the following characteristics was considered for the inclusion of articles: (i) articles focusing on the SARS-CoV-2 infection/COVID-19 disease; (ii) articles focusing on the impact on patients based in Italy or on the Italian hospital organization; (iii) articles detailing COVID-19-associated changes in the Italian hospital care for non-COVID-19 patients. All the investigated articles were published in English or Italian.

### Type of studies

Original papers, editorials, comments, research letters, case series and studies focusing on non-COVID patients in Italy were included.

### Data extraction and quality assessment

Data were extracted from the papers by one of the investigators (TP) and were subsequently checked for accuracy by other reviewers (GL, AnB). Disagreements regarding data extraction among reviewers were solved by consensus. Extracted data included: type of medical specialty involved (surgical, medical, or public health), geographical location (north, south, centre or nationwide), type of patients (COVID/non-COVID), type of study (article or research letter/comment/editorial). No quality assessment was possible as over 32% of entries were not articles but consisted of comments, research letters, opinions or editorials – for which no quality guidelines are available.

### Investigated Outcomes

This systematic review investigated the impact of the COVID-19 pandemic on patients’ healthcare provision and hospital organization in Italy. Our primary goal was to identify potential short-term and long-term effects on the health of non-COVID patients. Our secondary goals were to identify: (i) organizational and/or clinical settings and decisions that were particularly effective (or counterproductive) during the pandemic; and (ii) similarities and differences across medical specialties and regional areas.

## Results

The results are shown in Figure 1. After searching the databases, we identified a total of 4643 papers, from three different databases. Database merges and the removal of duplicate resulted in 1262 records, of which 100 were immediately removed as they were not related to COVID-19 (articles published before the pandemic in Italy). A total of 1162 records were then screened: 166 were removed as not relevant to Italy; 534 were removed as they referred to COVID-19 patients rather than non-COVID-19 patients. A total number of 247 were deemed eligible, of which 81 consisted of comments/letters/opinions/editorials.

**Figure 1.**
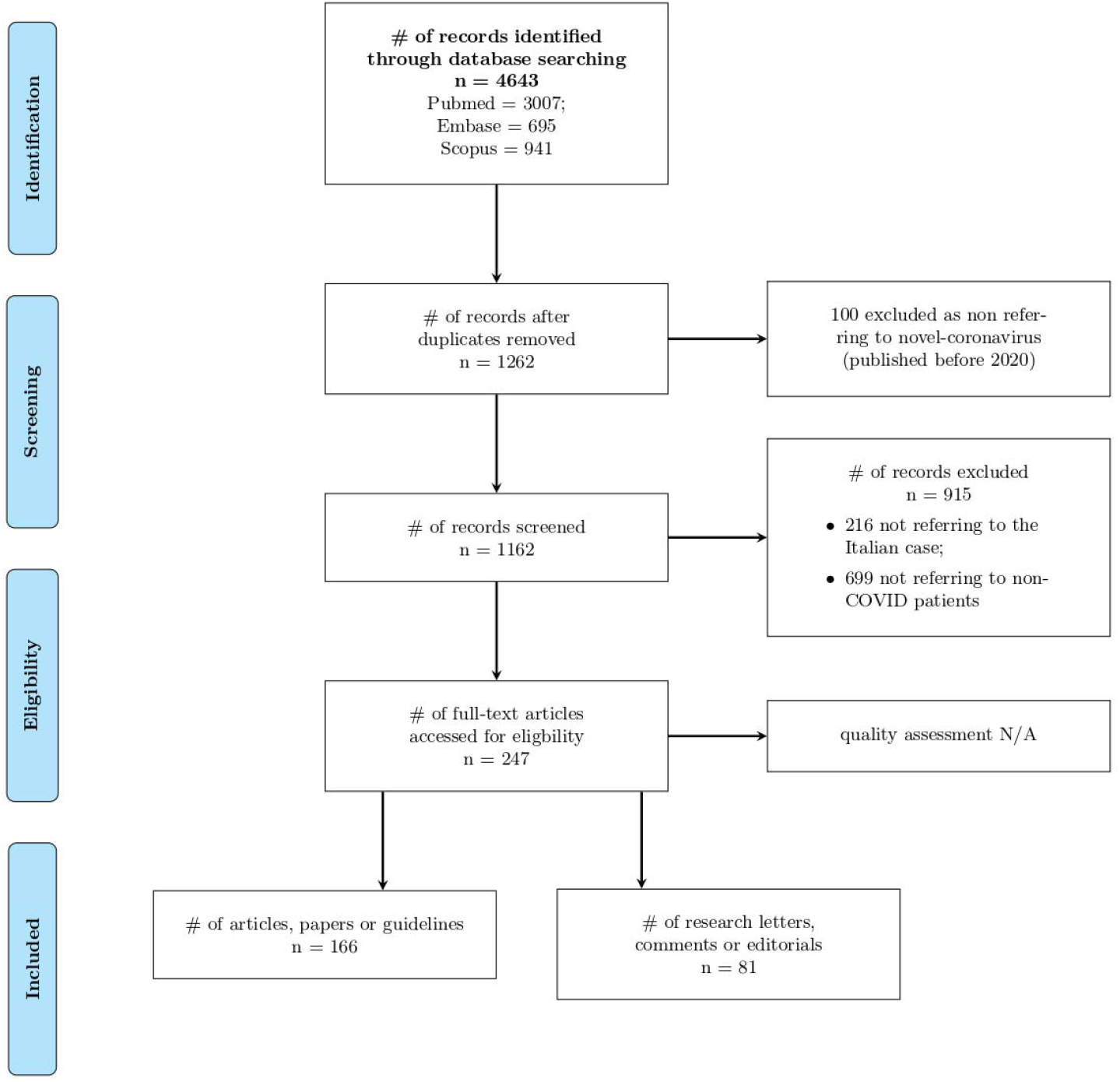
Results.

Oncology and radiotherapy were the most represented category (47 papers), followed by surgery (24 for general surgery, 9 for neurosurgery, 2 for cardio-surgery, 2 for vascular surgery, and 3 further types), cardiology (19) and dermatology 14. There was one paper each for rheumatology and microbiology.

Overall, 133 papers were related to clinical disciplines, 89 to surgery, and 24 to services. In terms of the geographical distribution, many papers provided general recommendations without a specific geographical identification (75). Lombardy was the most represented region (72), followed by Lazio (21) and Emilia Romagna (15). In the south, Campania was the most represented region (11), followed by Puglia (4). Marche, Piedmont and FVG had 7 papers each, Tuscany and Veneto 9 each. Overall, 73 were general/nationwide, 105 pertained to northern regions, 54 to central regions and 15 to southern regions and islands. The studies included are reported in Table 2, Supplementary Material.

**Table 2.**
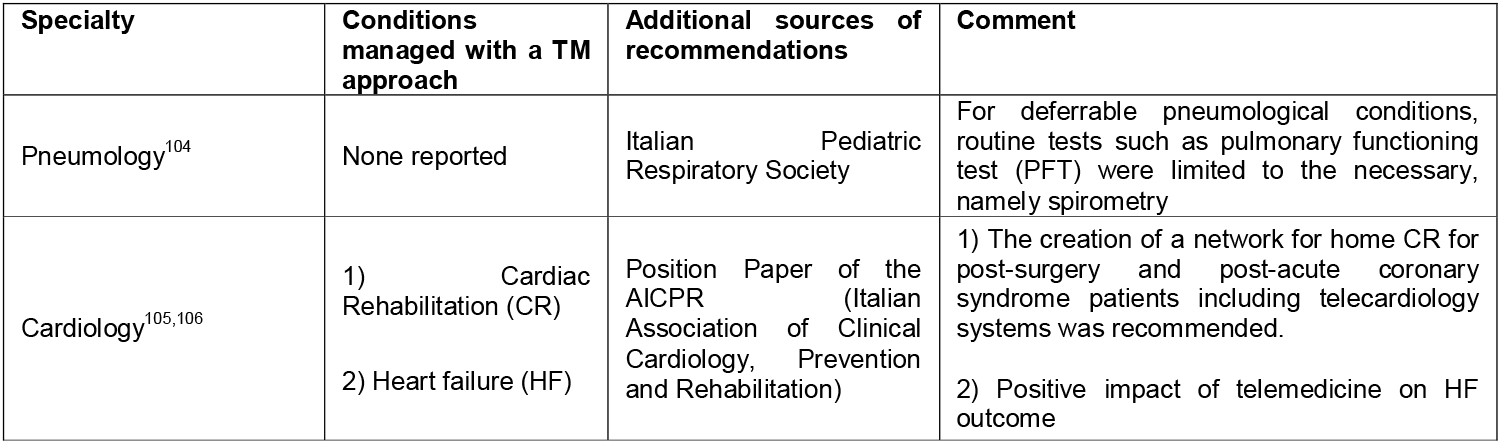
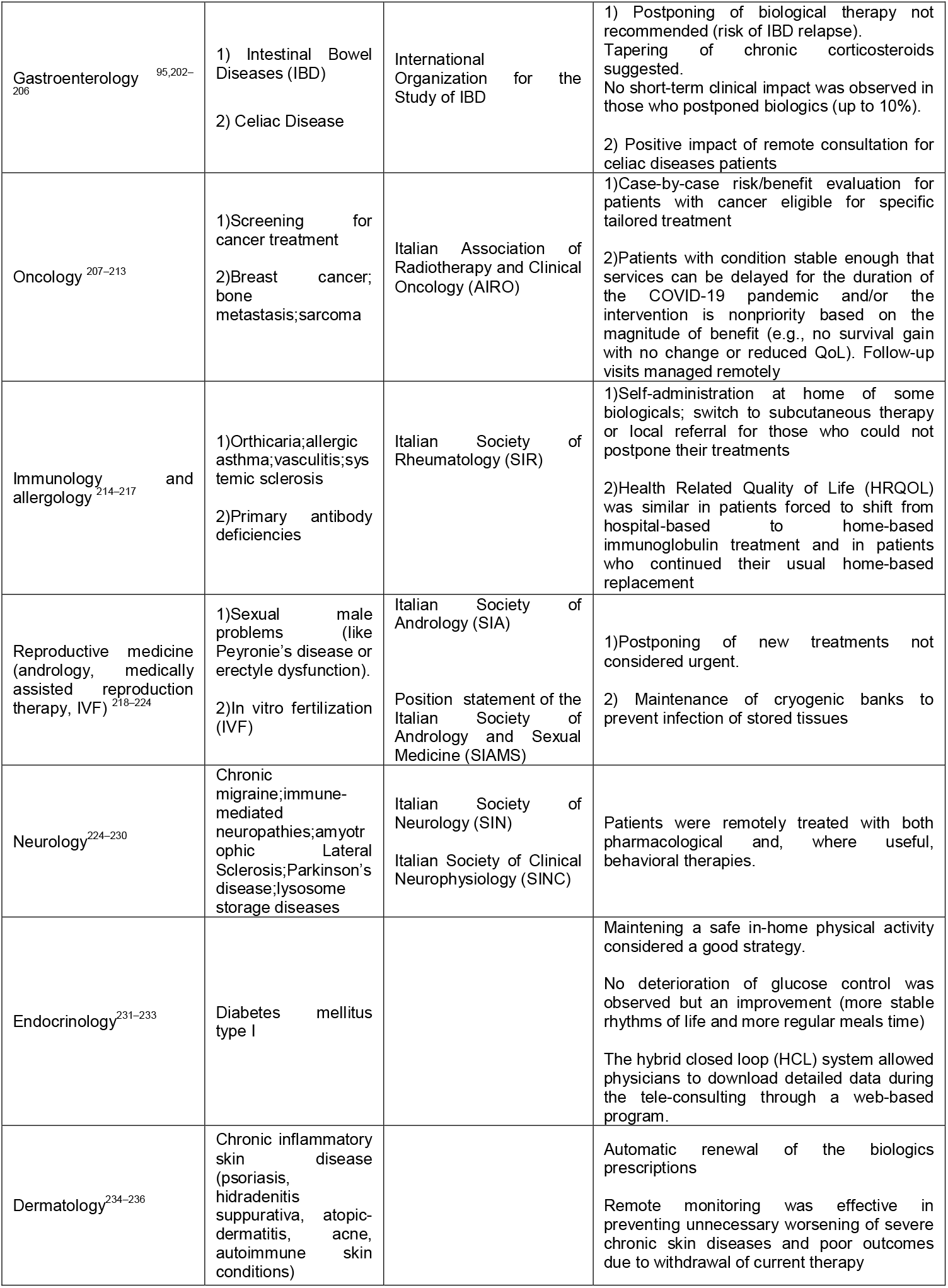
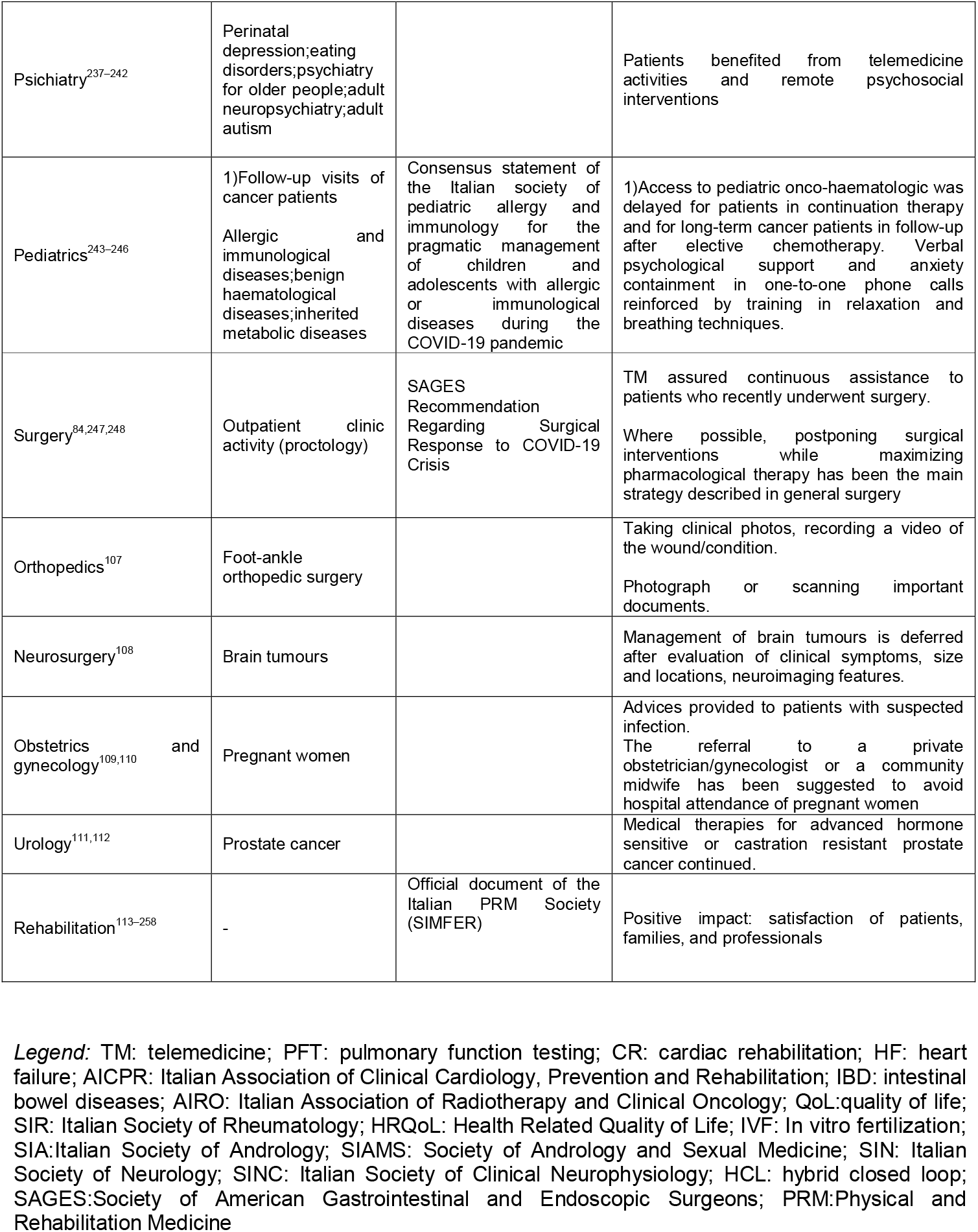
Management of deferrable conditions and telemedicine during COVID-19 pandemic in Italy.

| Specialty | Conditions managed with a TM approach | Additional sources of recommendations | Comment |
| --- | --- | --- | --- |
| Pneumology<sup>104</sup> | None reported | Italian Pediatric Respiratory Society | For deferrable pneumological conditions, routine tests such as pulmonary functioning test (PFT) were limited to the necessary, namely spirometry |
| Cardiology<sup>105,106</sup> | 1) Cardiac Rehabilitation (CR)<br>2) Heart failure (HF) | Position Paper of the AICPR (Italian Association of Clinical Cardiology, Prevention and Rehabilitation) | 1) The creation of a network for home CR for post-surgery and post-acute coronary syndrome patients including telecardiology systems was recommended.<br>2) Positive impact of telemedicine on HF outcome |
| Gastroenterology<sup>95,202–206</sup> | 1) Intestinal Bowel Diseases (IBD)<br>2) Celiac Disease | International Organization for the Study of IBD | 1) Postponing of biological therapy not recommended (risk of IBD relapse). Tapering of chronic corticosteroids suggested. No short-term clinical impact was observed in those who postponed biologics (up to 10%).<br>2) Positive impact of remote consultation for celiac diseases patients |
| Oncology<sup>207–213</sup> | 1) Screening for cancer treatment<br>2) Breast cancer; bone metastasis; sarcoma | Italian Association of Radiotherapy and Clinical Oncology (AIRO) | 1) Case-by-case risk/benefit evaluation for patients with cancer eligible for specific tailored treatment<br>2) Patients with condition stable enough that services can be delayed for the duration of the COVID-19 pandemic and/or the intervention is nonpriority based on the magnitude of benefit (e.g., no survival gain with no change or reduced QoL). Follow-up visits managed remotely |
| Immunology and allergology<sup>214–217</sup> | 1) Orticaria; allergic asthma; vasculitis; systemic sclerosis<br>2) Primary antibody deficiencies | Italian Society of Rheumatology (SIR) | 1) Self-administration at home of some biologics; switch to subcutaneous therapy or local referral for those who could not postpone their treatments<br>2) Health Related Quality of Life (HRQOL) was similar in patients forced to shift from hospital-based to home-based immunoglobulin treatment and in patients who continued their usual home-based replacement |
| Reproductive medicine (andrology, medically assisted reproduction therapy, IVF)<sup>218–224</sup> | 1) Sexual male problems (like Peyronie's disease or erectile dysfunction).<br>2) In vitro fertilization (IVF) | Italian Society of Andrology (SIA)<br>Position statement of the Italian Society of Andrology and Sexual Medicine (SIAMS) | 1) Postponing of new treatments not considered urgent.<br>2) Maintenance of cryogenic banks to prevent infection of stored tissues |
| Neurology<sup>224–230</sup> | Chronic migraine; immune-mediated neuropathies; amyotrophic Lateral Sclerosis; Parkinson's disease; lysosome storage diseases | Italian Society of Neurology (SIN)<br>Italian Society of Clinical Neurophysiology (SINC) | Patients were remotely treated with both pharmacological and, where useful, behavioral therapies. |
| Endocrinology<sup>231–233</sup> | Diabetes mellitus type I |  | Maintaining a safe in-home physical activity considered a good strategy.<br>No deterioration of glucose control was observed but an improvement (more stable rhythms of life and more regular meals time)<br>The hybrid closed loop (HCL) system allowed physicians to download detailed data during the tele-consulting through a web-based program. |
| Dermatology<sup>234–236</sup> | Chronic inflammatory skin disease (psoriasis, hidradenitis suppurativa, atopic dermatitis, acne, autoimmune skin conditions) |  | Automatic renewal of the biologics prescriptions<br>Remote monitoring was effective in preventing unnecessary worsening of severe chronic skin diseases and poor outcomes due to withdrawal of current therapy |
| Psichiatria<sup>237-242</sup> | Perinatal depression;eating disorders;psychiatry for older people;adult neuropsychiatry;adult autism |  | Patients benefited from telemedicine activities and remote psychosocial interventions |
| Pediatrics<sup>243-246</sup> | 1)Follow-up visits of cancer patients<br>Allergic and immunological diseases;benign haematological diseases;inherited metabolic diseases | Consensus statement of the Italian society of pediatric allergy and immunology for the pragmatic management of children and adolescents with allergic or immunological diseases during the COVID-19 pandemic | 1)Access to pediatric onco-haematologic was delayed for patients in continuation therapy and for long-term cancer patients in follow-up after elective chemotherapy. Verbal psychological support and anxiety containment in one-to-one phone calls reinforced by training in relaxation and breathing techniques. |
| Surgery<sup>84,247,248</sup> | Outpatient clinic activity (proctology) | SAGES Recommendation Regarding Surgical Response to COVID-19 Crisis | TM assured continuous assistance to patients who recently underwent surgery.<br>Where possible, postponing surgical interventions while maximizing pharmacological therapy has been the main strategy described in general surgery |
| Orthopedics<sup>107</sup> | Foot-ankle orthopedic surgery |  | Taking clinical photos, recording a video of the wound/condition.<br>Photograph or scanning important documents. |
| Neurosurgery<sup>108</sup> | Brain tumours |  | Management of brain tumours is deferred after evaluation of clinical symptoms, size and locations, neuroimaging features. |
| Obstetrics and gynecology<sup>109,110</sup> | Pregnant women |  | Advices provided to patients with suspected infection.<br>The referral to a private obstetrician/gynecologist or a community midwife has been suggested to avoid hospital attendance of pregnant women |
| Urology<sup>111,112</sup> | Prostate cancer |  | Medical therapies for advanced hormone sensitive or castration resistant prostate cancer continued. |
| Rehabilitation<sup>113-258</sup> | - | Official document of the Italian PRM Society (SIMFER) | Positive impact: satisfaction of patients, families, and professionals |

**Legend:** TM: telemedicine; PFT: pulmonary function testing; CR: cardiac rehabilitation; HF: heart failure; AICPR: Italian Association of Clinical Cardiology, Prevention and Rehabilitation; IBD: intestinal bowel diseases; AIRO: Italian Association of Radiotherapy and Clinical Oncology; QoL:quality of life; SIR: Italian Society of Rheumatology; HRQoL: Health Related Quality of Life; IVF: In vitro fertilization; SIA:Italian Society of Andrology; SIAMS: Society of Andrology and Sexual Medicine; SIN: Italian Society of Neurology; SINC: Italian Society of Clinical Neurophysiology; HCL: hybrid closed loop; SAGES:Society of American Gastrointestinal and Endoscopic Surgeons; PRM:Physical and Rehabilitation Medicine

### Management of emergencies

In general, non-COVID patients admitted to emergency departments (ED) decreased and remained well below the standard levels^7–11^.The youngest age classes declined dramatically, whilst the oldest age classes progressively increased, remaining considerably above the standard rate of the local ED.^8^

Table 1 shows the most relevant data regarding cardiovascular emergencies, including stroke. In northern Italy, the emergency gradually took over most of cardiology intensive care units (ICUs).^9^ As expected, the net effect of this reorganization was a significant reduction in sites and staff committed to the treatment of cardiovascular diseases.^10–11^ Comparing out-of-hospital cardiac arrests in the same period of the previous year, Baldi *et al*. found that the median arrival time of the emergency medical service was three minutes longer in 2020, and the proportion of patients who received cardiopulmonary resuscitation from bystanders was 15.6% lower. Among patients in whom resuscitation was attempted by emergency medical services, the incidence of out-of-hospital death was 14.9% higher in 2020 than in 2019.^7^ This finding was confirmed by additional studies highlighting an unpredictable decrease in acute coronary syndrome-related hospitalization in high-volume centres^12,13,14,15^ and pacemaker implantation during the weeks following the COVID-19 outbreak.^21–18^

Of note, one study showed a 49% reduction in acute heart failure admission to hospital.^19^ The patients admitted had longer door-to-balloon and symptoms to PCI times, higher hs-cTnI levels at presentation, lower residual LV function at discharge, and higher predicted late cardiovascular mortality on the GRACE score.^13^

A lower number of patients with minor strokes and TIAs, longer onset-to-door and door-to-treatment times for major strokes, and reduced number of transfers from spokes centres were also reported.^11,20^ As a result, the number of patients who underwent intravenous thrombolysis or bridging therapy (combined intravenous and thrombectomy) decreased by −□26% and −30%, respectively.

In addition, as a consequence of the reduction in the patient eligibility for bridging therapy, the number of primary thrombectomies (performed with all the necessary personal protective equipment owing to the risk of infection^21^) increased by 41%. Most of these patients had very serious strokes that would have benefited from early diagnoses.^22^

The decrease in hospital admissions, confirmed by a survey across multiple countries including Italy^23,24^, resulted in increased door-to-needle times and therapeutic windows being missed for patients suffering from severe strokes.^25^ More studies are needed to confirm these data and to evaluate the impact on death rates; however, recent reports from other countries seem to confirm this trend.^26,27^

The reduction in available ICU beds, massively dedicated to COVID-19 patients with acute respiratory failure, and the fear of infection resulted in the shrinkage of surgical activities in all fields^28–38^ and a reduction in urgent endoscopic procedures in COVID-free hospitals.^29^

Each sub-specialty defined various non-deferrable surgical procedures that had to be guaranteed, causing a drop in consultations requested by emergency departments as in the case of urgent urology.^30^ The surgical community were also faced with a shortage of blood components, derived from fewer donations due to lockdown and fear of infection.^31^ To compensate for the initial fall (−10%) in blood donations in the first week of March, the government promoted a national media campaign on the importance and safety of blood donation as a priority to maintain basic healthcare services for non-COVID patients.^32^ No blood-transmitted SARS-CoV-2 infection has been reported to date.^33^

Children’s emergency departments also showed a substantial decrease in visits.^34^ This might reflect the scarcity of resources or the reluctance of parents and health care workers to expose children to the risk of viral infection in a health-care setting, in addition to lower rates of acute infections and trauma.^35^ However, this phenomenon has been detrimental to the health of non-COVID child patients: 12 cases of delayed access to hospital care were reported during the week March 23– 27 across five hospitals of an Italian Children’s Hospital Research Network. Half of the children were admitted to an ICU and four died, highlighting the high risk of delaying access to hospital care.^36^ As a result, life-threatening conditions (i.e. abdominal pain, severe ketoacidosis) seemed to be more frequent, requiring in some cases an aggressive approach.^35^

The same phenomenon affected dermatology^37^. Tartari et al. compared two different weeks, before and after the outbreak of the COVID-19 pandemic in Italy, showing a decrease in unjustified referrals (93% reduction) in dermatological emergency services.^38^

Despite medical care for emergencies and urgent treatments being continuously provided throughout the pandemic, the lack of personnel, resources, and ICUs beds along with the patients’ fear of being infected in hospital affected patient management and substantially delayed the provision of ordinary medical activities. These initial data seem to show a decrease in emergencies and an increase in mortality. We strongly suggest undertaking detailed studies to further explore this phenomenon.

### Management of non-deferrable conditions: the hub-and-spoke system

To tackle the massive impact of the overflow of SARS-CoV-2 infected patients, hospitals in Italy had to undergo a significant reorganization.^39–40^ In order to manage conditions needing non-deferrable treatment while avoiding the risk of infection, hub-and-spoke centres were created and widely used throughout the country.^50,55–58^ In the hub-and-spoke model, a main campus or hub supplies the most intensive medical services, while satellite campuses or spokes offer more limited services at sites distributed across the neighbouring area.^41^

Neurological surgery was particularly affected by the ICU reorganization, as it often requires a period of intensive monitoring in ICUs.^22,60–62^ All the cases of elective neurological surgery were deferred, while urgent neurosurgical pathologies (above all traumas) and non-deferrable tumour cases were transferred to hubs.^27,29–33,61,67,68^ A specific urgency classification for tumour cases was developed based on symptoms, radiological findings, and suspected malignancy of the lesion.^16,42–44^ Some minimal activities were still performed at spoke centres, only for very critical cases or when specific tools were required (i.e. gamma knife treatment of neoplastic lesions).^16,42,45,46^

This provided an unprecedented opportunity for transversal collaboration among different teams, representing real innovation in such a competitive setting.^58,64^

A hub-and-spoke system was also organized for vascular surgery and cardiac surgery units. All elective surgery was reduced and urgent surgery (including aortic aneurysms, valvular diseases or severe coronary diseases) was performed only in hub centres, preferring the endovascular to the open surgical approach whenever possible.^70–72^ A transcatheter approach was generally preferred, as it usually does not require an ICU bed or a ventilator.^47^

The limitation of all non-urgent surgical activities also applied to general surgery^73–78^ and obstetrics.^48,49^ In highly-infected areas (such as Lombardy), hub centres were created^50^ to treat only advanced symptomatic tumours^81–83^, while elective oncological surgery procedures continued to be performed in less affected regions.^50,51^ This was an important issue for oncological patients, especially older ones.^85–88^

Many possible ways of minimizing the risks were proposed: to postpone treatments or elective surgery for stable cancer in endemic areas, to provide patients with greater personal protection, and to offer more intensive surveillance or treatment.^52–54^ For example, neoadjuvant treatments were recommended or increased^55^ to defer surgical admission for as long as possible.^50,51,55,56^ For other medical conditions requiring surgery under particular circumstances, such as relapse of inflammatory bowel disease, dedicated hubs were identified.^94–57^

Interestingly, a tendency toward treatments aimed at reducing hospitalization was also found in medical oncology.^58,59^

Some regions such as Tuscany created protocols for home care in order to avoid exposure to hospital settings.^60^ Oncological care delivery and cancer diagnosis^100–102^ were dramatically reduced by the SARS-CoV-2 outbreak, even though suboptimal care and treatments may result in worse cancer-related outcomes. Oncologists were thus asked to preserve the continuum of care of patients, while adopting mitigation strategies to reduce the likelihood of infection in all cancer patients.^103–61^

Arduino et al. described a worrying delay in the diagnosis of oral cancer in north-west Italy during the Covid pandemic.^62^ Moreover, the cessation of elective activities, screening programs^63^, and the drastic reduction in services regarding breast cancer restricted evaluations to only clinical observations of palpable lesions with the elevated risk of missing new diagnoses.^64^, Although not requiring a structural reorganization, palliative care was forced to find a new balance between family member visits and patients’ needs.^116–119^

In orthopaedics, the reorganization led to the identification of poly-specialist major trauma centres and specialistic referral centres for minor trauma or non-deferrable orthopedic surgeries (i.e. septic arthritis or malignant tumours).^120–127^ There was a reduction in the number of proximal femur fractures in two centres^65^, as well as a reduction in hip and knee arthroplasties.^66^

A similar re-organization was also carried out for plastic surgery: only post-traumatic, oncological and burn treatments were guaranteed.^130–132^ A new approach based on enzymatic debridement was proposed for burns in order to reduce the need for burn surgery.^67^

In urology, only urgent, non-deferrable procedures (colicky flank pain, gross hematuria and acute urinary retention) were authorized after careful multidisciplinary evaluation,^134–138^ which led to a drop in urological surgical activities.^68^ Whenever possible, alternative treatments not requiring general anaesthesia (i.e. radiotherapy for genitourinary cancers) were suggested as being preferable.^69,70,71^

Oral and maxillofacial surgery^72^, otolaryngology^73,74^ and ophthalmology^144–147^ also suspended all non-urgent treatments, especially considering the high risk of infection of healthcare workers while manipulating the upper airways and eyes.^75^ Only the treatment of trauma, malignant neoplasms, and severe infections was guaranteed.^142,149–152^

In the context of radiotherapy, all follow-up visits involved a phone call in advance in order to postpone non-urgent cases.^153–156^ The initial consultations of patients needing treatment for malignant tumours were conducted as normal^157–159^, as were certain treatments such as bone metastase radiotherapy.^76^ Specific approaches, such as short fractionated radiotherapy, were suggested.^77^

All non-urgent and deferrable radiation treatments were delayed, while therapies for patients with a better prognosis (benign and functional diseases) were postponed.^78,79^

Dermatology departments were also involved in an extensive reorganization.^80,81^ Dermatological anti-neoplastic treatments were provided in the dermatology clinics of many centres, such as Bologna^82^, Naples^83^, Modena^84^ and Ancona^85^, which also maintained urgent dermatological procedures and consultations required by other hospital wards. As awareness of the severity of the COVID-19 increased, some patients were concerned about continuing their medications, however all centres followed specific recommendations and advised patients not to suspend these drugs without consultation.^170–173^

Lastly, microbiology labs underwent unprecedentedly high workloads with the increasing number of samples (swabs or serological tests) to analyze for the identification of Sar-Cov-2 infection. An extensive reorganization of the microbiology lab activities thus also occurred. In a large teaching hospital in Rome, the introduction of night shifts and the creation of a dedicated team significantly improved the number of samples processed, without interfering with the daily laboratory routines.^86^

Hub-and-spoke centres represented an important change in care provision, especially in the most affected regions involving almost all specialties. However, data on the efficacy of this reorganization, measured in terms of health outcomes (such as mortality) are lacking. To date, only a few reports^100,102,113^ suggest a tendency toward a decrease in diagnosis for non-deferrable conditions despite the hub-and-spoke organization. Further epidemiological studies are needed to verify the emerging picture and to evaluate the impact on mortality rates.

### Replacement therapies: dialysis and transplantation

Dialysis units experienced a profound change in their management, with the introduction of COVID-19 isolation rooms and the identification of dedicated healthcare professionals.^87,88^ Rombolà et al. proposed three actions to be taken in order to dialyze non-COVID patients safely: hygiene measures, the use of PPE to protect patients and the healthcare team, and the protection of the dialysis ward with an isolated area for testing patients suspected of infection.^89^

In general, all transplant programs were profoundly affected by the pandemic.^178–180^ First of all, the wall-to-wall screening of donors and recipients was established in order to identify positive patients that would not be able to donate or receive blood, in view of the high mortality rates of COVID-19 in immunocompromised patients.^78,180– 183^ Secondly, the widespread reduction in available ICU beds led to an estimated 15% drop in transplants compared with the average of the last five years^90^, such as liver transplantations.^91^ Transplantation was thus suggested only in true end-stage organ failure, preferring conservative treatments (maximizing pharmacological therapy) in all other patients.^184,185,187^

### Management of deferrable conditions and telemedicine

The management of chronic conditions also suffered.^92^ Cesari et al. found that the integration of care services collapsed: admissions to post-acute/long-term care facilities were reduced and several person-tailored interventions were suspended – e.g., physical therapists for mobilization.^93^ Lasevoli et al. reinforced the view that the current pandemic has had dramatic consequences for the mental health of serious psychiatric patients.^94^ All this inevitably led to a drastic reduction and a substantial reorganization of the clinical activity in many specialties^95^ (Table 1), postponing elective treatments and switching to telemedicine (TM)^96^ for consultation or not to leave vulnerable high-need patients without proper follow-up^228,234^.

The implementation of TM occurred in different ways and to varying degrees depending on the specific center and specialty. An online questionnaire administered to the 176 Directors of Italian Radiation Oncology Departments revealed that, to guarantee the continuity of care, telematic consultations for RT treatments were activated in 78 centres (62.4%)^155^. A similar survey for RT centres in Lombardy region revealed that 84% of RT facilities cancelled out-patient follow-up visits, 68% activated telematic consultation and 30% adopted working from home solutions^156^. Another survey administered to 122 medical oncology departments homogeneously distributed on the national territory, revealed that in 72% of cases alternative ways to get in touch with patients have been used, like telephonic interviews with interpretation of laboratory and radiologic examination reports. According to Pietrantonio et al., WhatsApp turned out to be adequate to give a rapid answer to most queries from oncologic patients^108^. Brunasso et al. started a teledermatologic service in smartworking using phone calls and emails by which could monitor almost 94% of their patients^234^. In a Department of Urology in Northern Italy, 55% cases were screened undergoing telephone consultation^253^.

TM has been shown to have beneficial impacts on heart failure outcomes in a comparative analysis between 2020 and 2019 by Salzano et al.^200^. Finally, TM positively impacted patients’ life as documented by a survey in which 85% of patients were satisfied with the remote interview modality and the reduction of economic and time costs related to going to the clinic. Most of those subjects (90%) expressed their willingness to continue to be included in remote evaluation programs^228^.

TM consists in the distribution of health-related services and information via telecommunication technologies and represents an elective approach to managing deferrable conditions during the COVID-19 pandemic in Italy. Almost all specialties benefited from TM during the pandemic. In many cases, the short-term results have been particularly encouraging, although a longer follow-up is needed to assess the efficacy of these measures. The results are summarized in Table 2.

TM has deeply influenced non-COVID patient care, enabling the remote diagnosis and monitoring of patients and allowing clinical data sharing between patients and physicians. However, the long-term effects on common health outcomes need to be carefully studied.^97,98^ Avoiding face-to-face contact via TM has been one of the most effective measures to limit the spread of SARS-Cov-2 infection, although many issues have been raised, such as privacy management and the lack of clear guidelines.^194–198^

## Discussion

On 11 March 2020, the World Health Organisation (WHO) declared COVID-19 to be a pandemic.^1^ However, Italy was already in lockdown, with decrees aimed at limiting mobility and strengthening the National Health System. On March 9, 2020^99^, most outpatient services were temporarily suspended, except for a few treatments that were considered urgent and non-deferrable. Clinical support for early isolation, treatment, and, where needed, intensive care of COVID-19 patients (or suspect cases) became the priority, with a massive allocation of dedicated resources.

A large increase in all-cause mortality was revealed during the epidemic, greater than the number of deaths attributed to COVID-19 cases. The possible causes of this increase include the large number of severe undiagnosed COVID-19 cases, the reduced access to health services due to the disruption of normal working processes or due to the fear of contamination of sick patients affected by other diseases, and possibly other factors.^100^

We provided a snapshot, across all medical specialties, of how the provision of treatments to non-COVID patients in Italy has been impacted by the shortage of resources imposed by the pandemic.

Measures put in place to mitigate the outbreak, such as social distancing and confinement, contributed to discourage access to emergency department (ED) all over the country, also for those conditions requiring urgent care. As a result, there was a significant decrease in overall ED admissions and a substantial reduction in all-specialty surgical consultations.^9-11^ Cardiovascular emergencies payed significant tolls with a significant delay in time-sensitive emergency operations.^9,19^ More recent evidences, consistent with our results, showed a significant decrease in mean number of endovascular therapies per hospital performed before and after COVID-19 confinement along with a significant increase in mean stroke onset-to groin puncture time^260^.A delayed presentation of STEMI patients that may led to worsened prognosis and unnecessary deaths has also been observed^261^. Moreover, an additional study confirmed that more in-hospital cardiovascular deaths occurred in March 2020 compared with March 2019, a finding possibly due to late hospital presentations and consequent greater disease severity that affected eligibility and outcome of cardiovascular procedures^262^.

As stated before, the hospitalization system was remodelled to allocate appropriate resources to manage patients with COVID-19; consequently, hubs were identified for specialized medical activities. Hub-and-spoke centres represented an important change in care provision, especially in the most affected regions, involving almost all specialties. However, data on the efficacy of this reorganization, measured in terms of health outcomes (such as mortality) are lacking. To date, only few reports^100,102,113^ suggest a tendency toward a decrease in diagnosis for non-deferrable conditions despite the hub-and-spoke organization. Cautious and evidence-based studies are needed to properly assess the overall impact of this model on measurable outcomes. The hub-and-spoke system seems to be a valid model, at least, in the management of ischemic emergencies^264^; further epidemiological studies are needed to verify the emerging picture and to evaluate the impact on mortality rates.

A pandemic is a dynamic scenario, requiring reorganization and flexibility of the healthcare delivery. TM, which consists in the distribution of health-related services and information via telecommunication technologies, proved a pragmatic approach to managing deferrable conditions during the COVID-19 pandemic in Italy. TM has deeply influenced non-COVID patient care, enabling the remote diagnosis and monitoring of patients and clinical data sharing between patients and physicians with adequate protection for both. Moreover, TM allows for more flexibility on the side of both the clinician and the patient, as consultations can easily be rescheduled, and meetings can be held from home^234^.

Almost all specialties benefited from TM during the pandemic, with short-term results particularly encouraging in some cases. In general, the pandemic has demonstrated that information technologies should be more promoted independently from this specific contex^155^. However, a longer follow-up is needed to assess the efficacy of these measures on common health outcomes^97,98^. Avoiding face-to-face contact via TM has been one of the most effective measures to limit the spread of SARS-Cov-2 infection, although many issues have been raised, such as privacy management and the lack of clear guidelines.^194–198^ We strongly encourage to overcome these limitations to further promote the multiple opportunities of TM in tune with its pivotal role during the second phase of COVID-19 pandemic in Italy^263^.

One limitation of this systematic review is the heterogeneity in publication type, which prevented the execution of a meta-analysis to summarize the findings together with a quality assessment. Another important issue is the potential underreporting, although the studies did cover experiences from the whole country.

Our work thus suggests that a public health crisis has resulted from the pandemic, a concern raised in other countries too, such as France.^259^ More detailed, population-based studies are needed to assess whether emergency management benefited from the reorganization adopted. Nevertheless, the hub-and-spoke system and telemedicine undoubtedly played – and continue to play – a crucial role in dealing with non-deferrable and deferrable conditions, respectively.

## Data Availability

The authors confirm that the data supporting the findings of this study are available within the article [and/or] its supplementary materials.

## Contributions

T.P. and G.L. conceived and designed the study. T.P. collected the data. G.L, M.M.O, An.B, G.A., Al.B. analyzed and selected the data. F.C. and T.P. supervised data analysis. G.L, M.M.O., An.B, G.A., Al.B., T.P., wrote the manuscript. M.M.O. and An.B. revised and edited the manuscript. L.Z. and P.R. revised the final version of manuscript.

## Acknowledgments

The work was supported by the Center for Excellence and Transdisciplinary Studies (CEST) and CRT Foundation, Turin. The funder of the study had no role in the study design, data collection, data analysis, data interpretation, or writing the report. All authors had full access to all study data and had final responsibility for the decision to submit for publication. The authors are thankful to Professor Paolo Vineis for his critical assistance in the writing of the paper.

## Supplementary Material

**Table 1.**
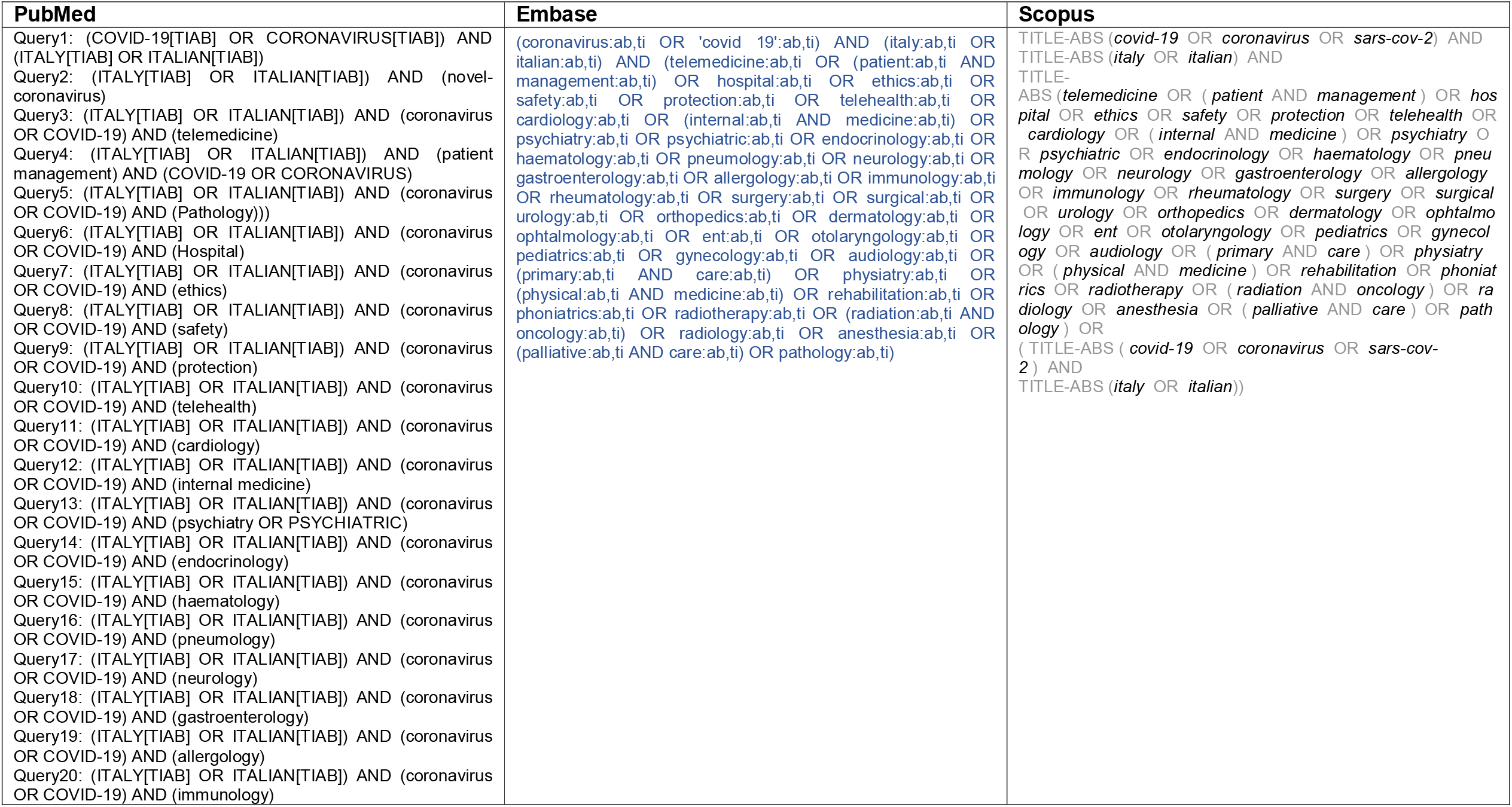
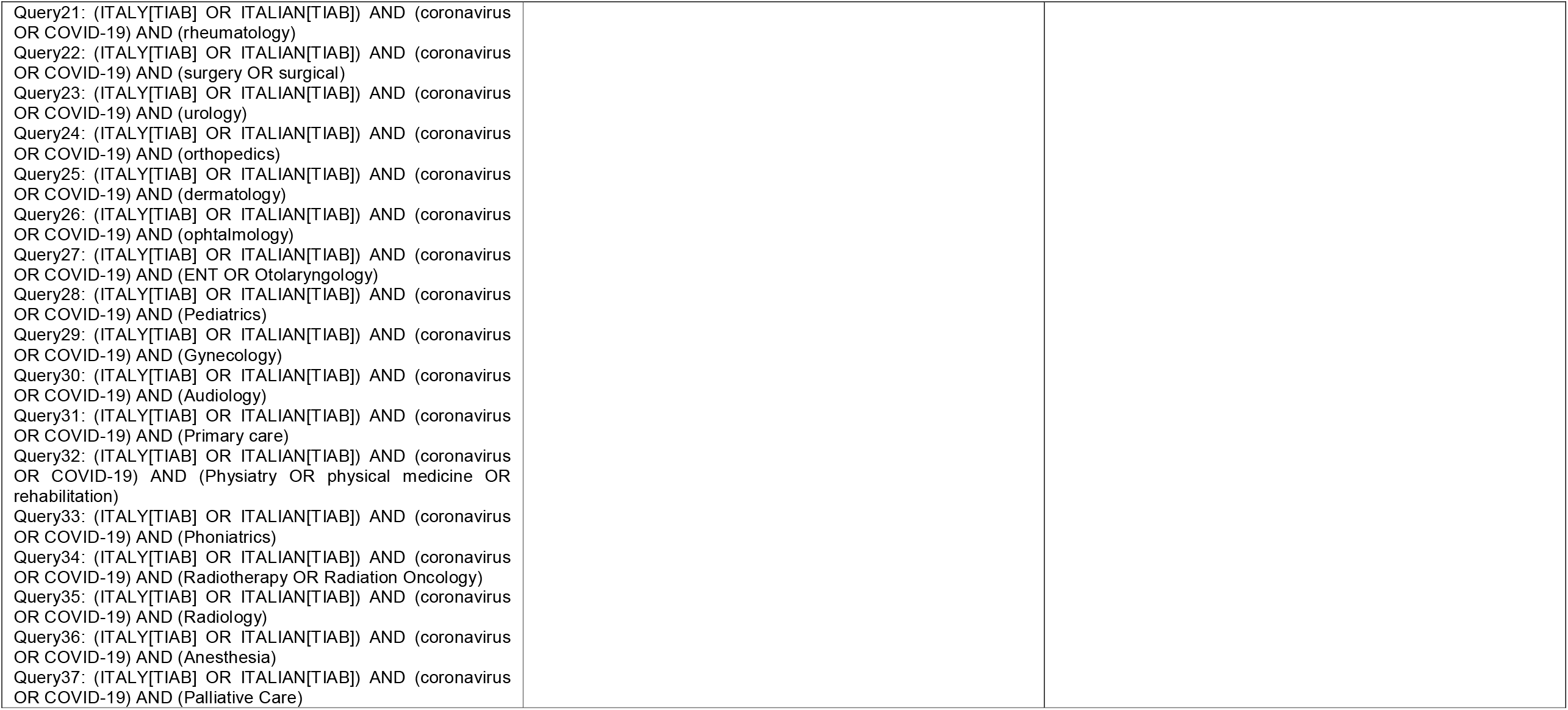
Full search strategy adopted in three different databases.

**Table 2.**
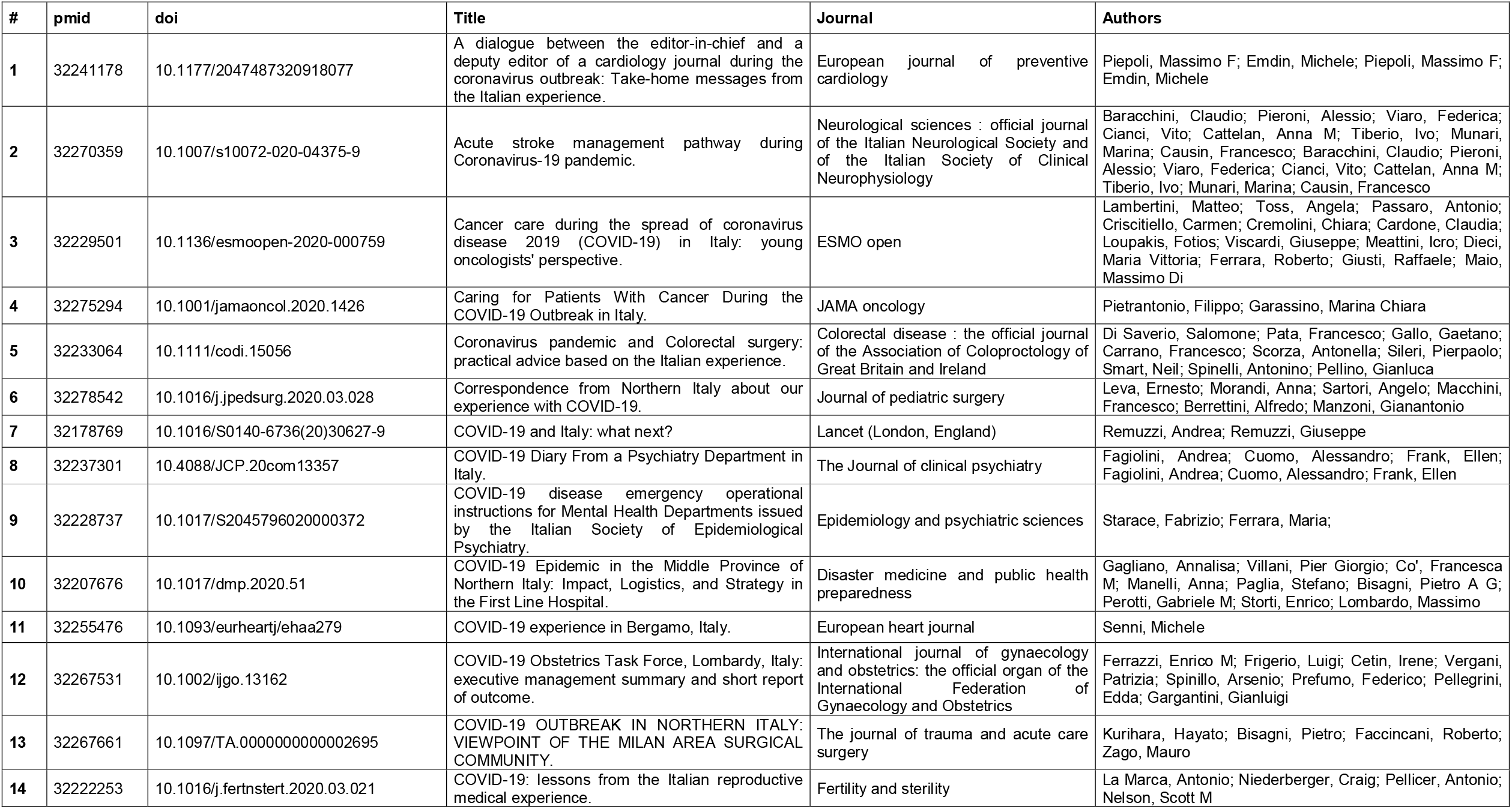
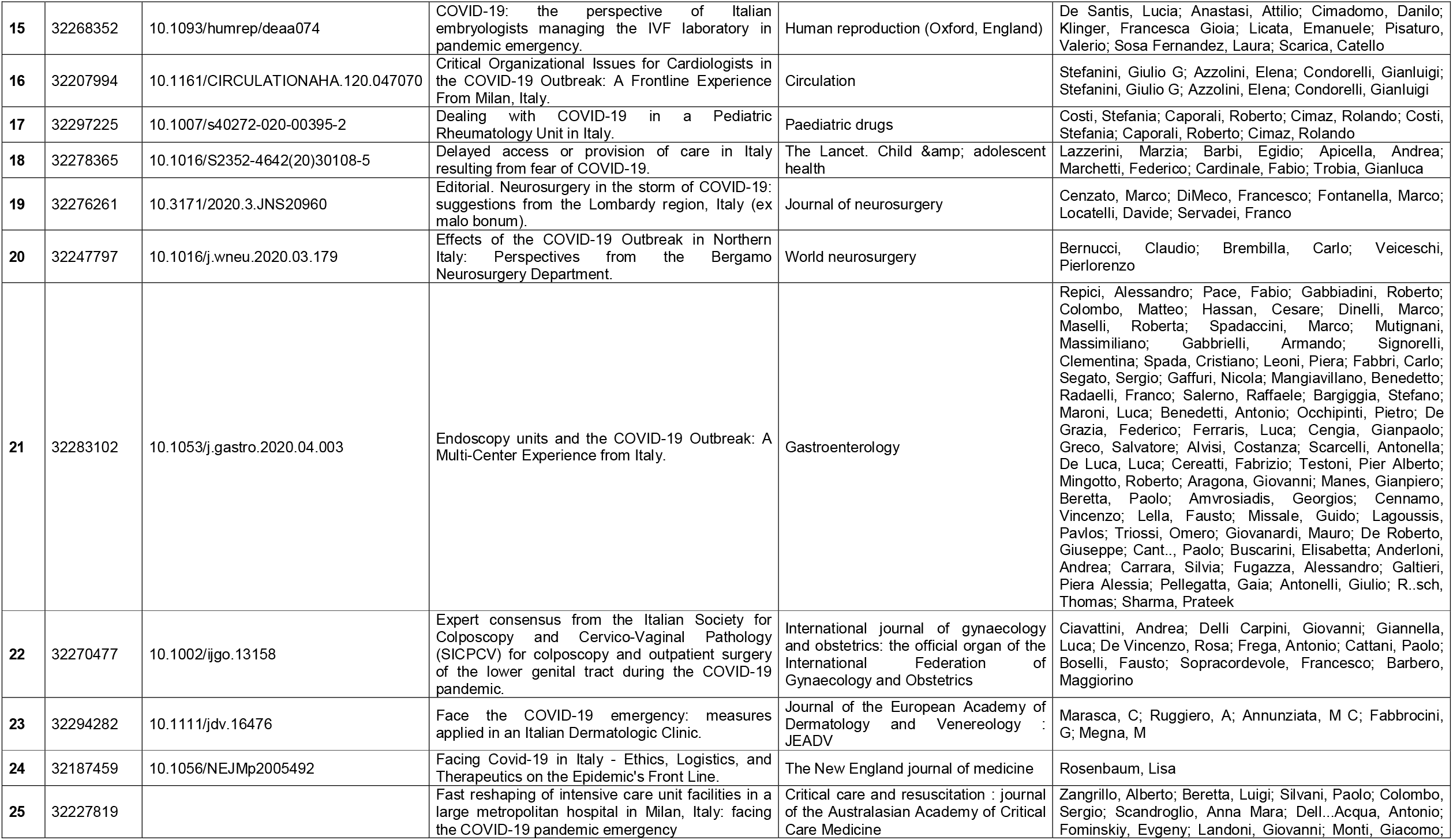
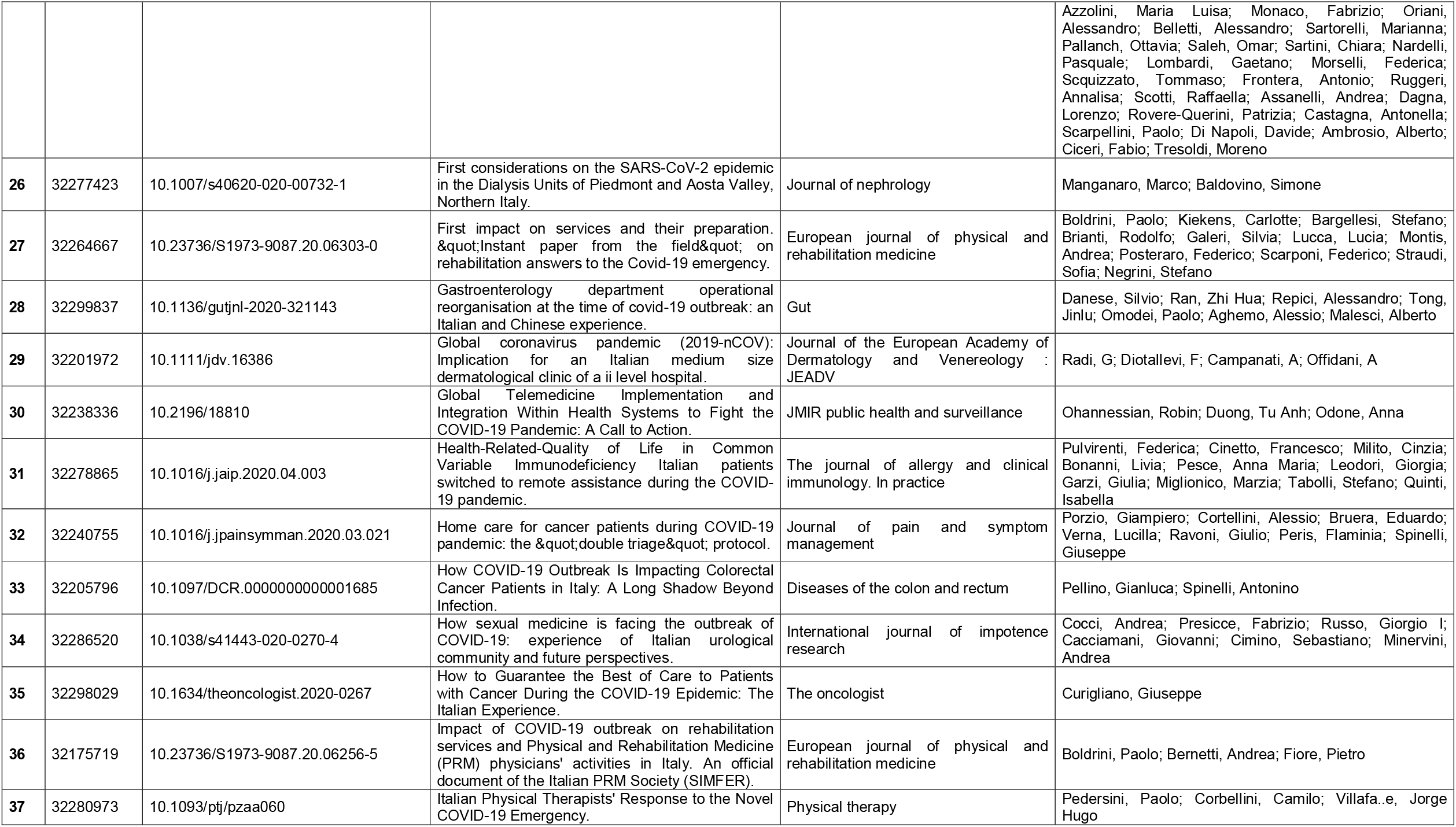
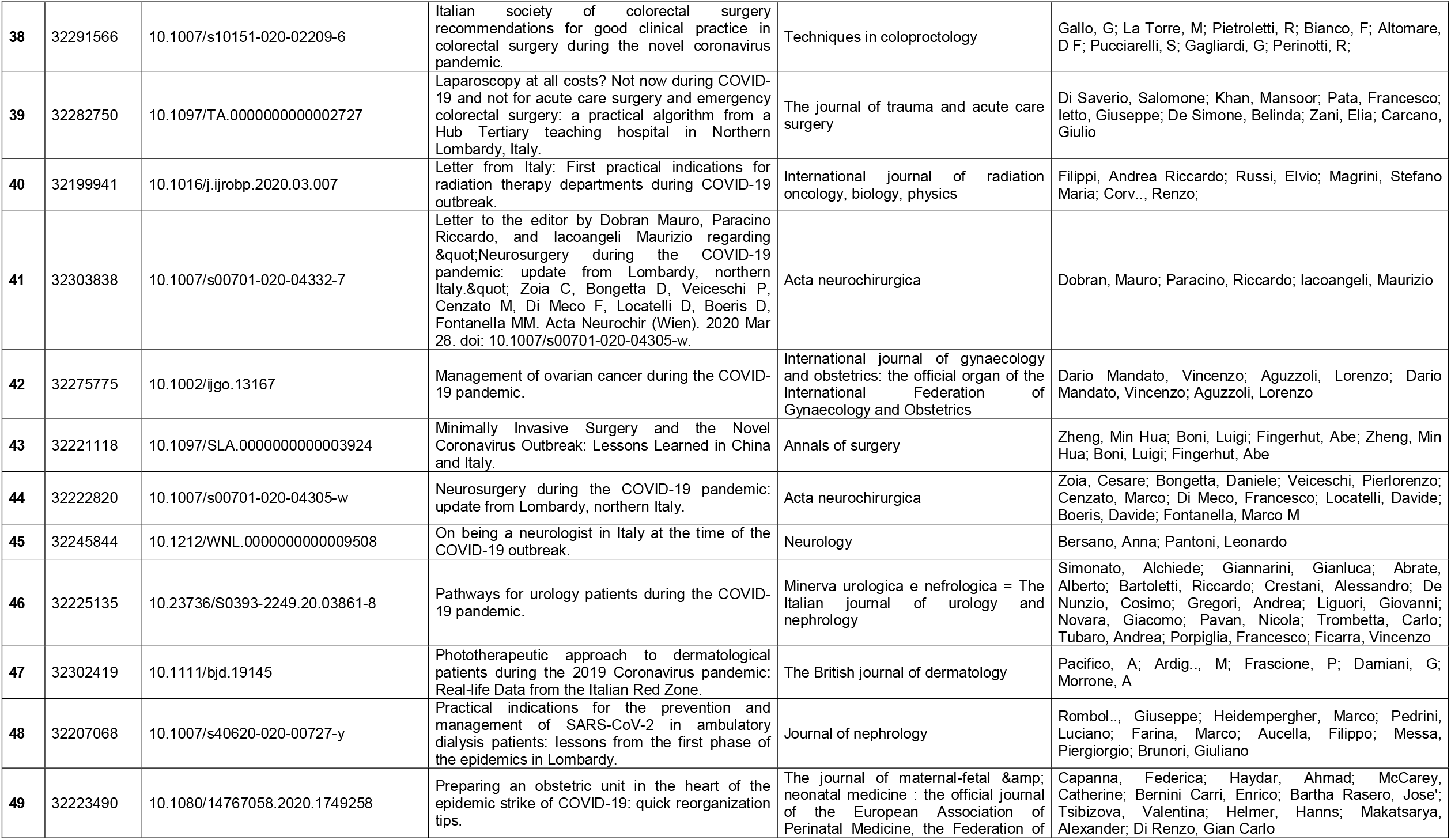
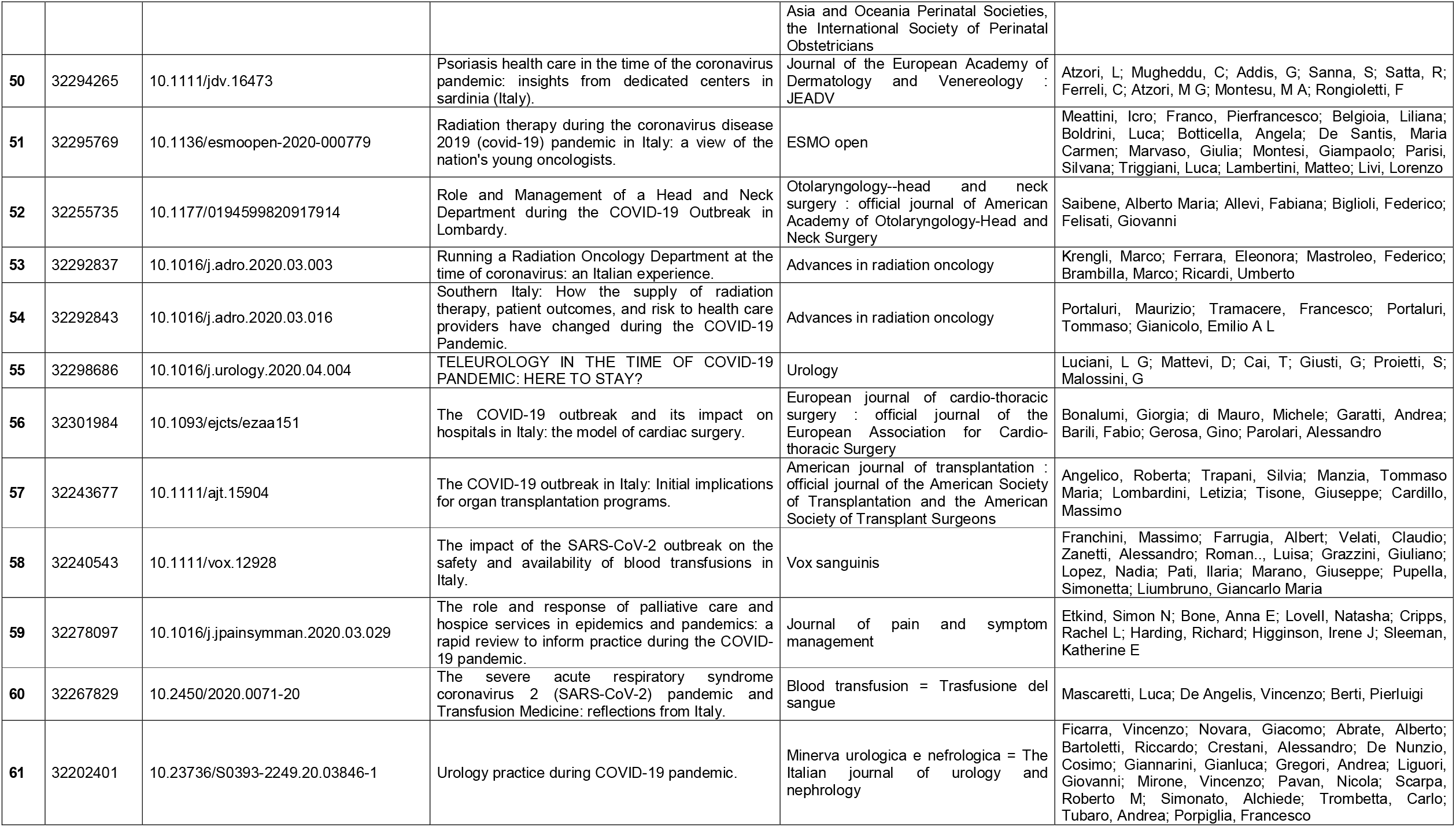
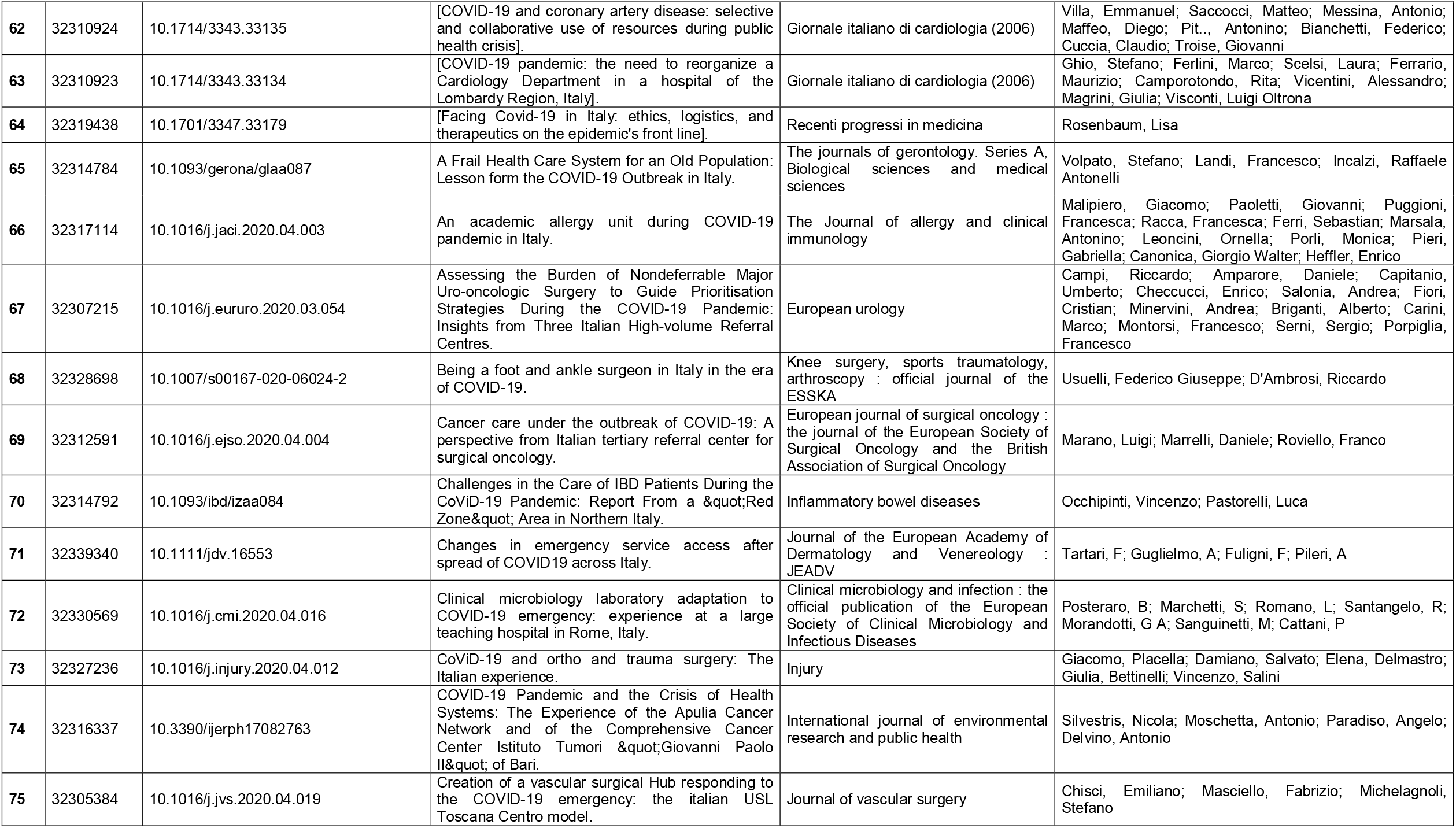
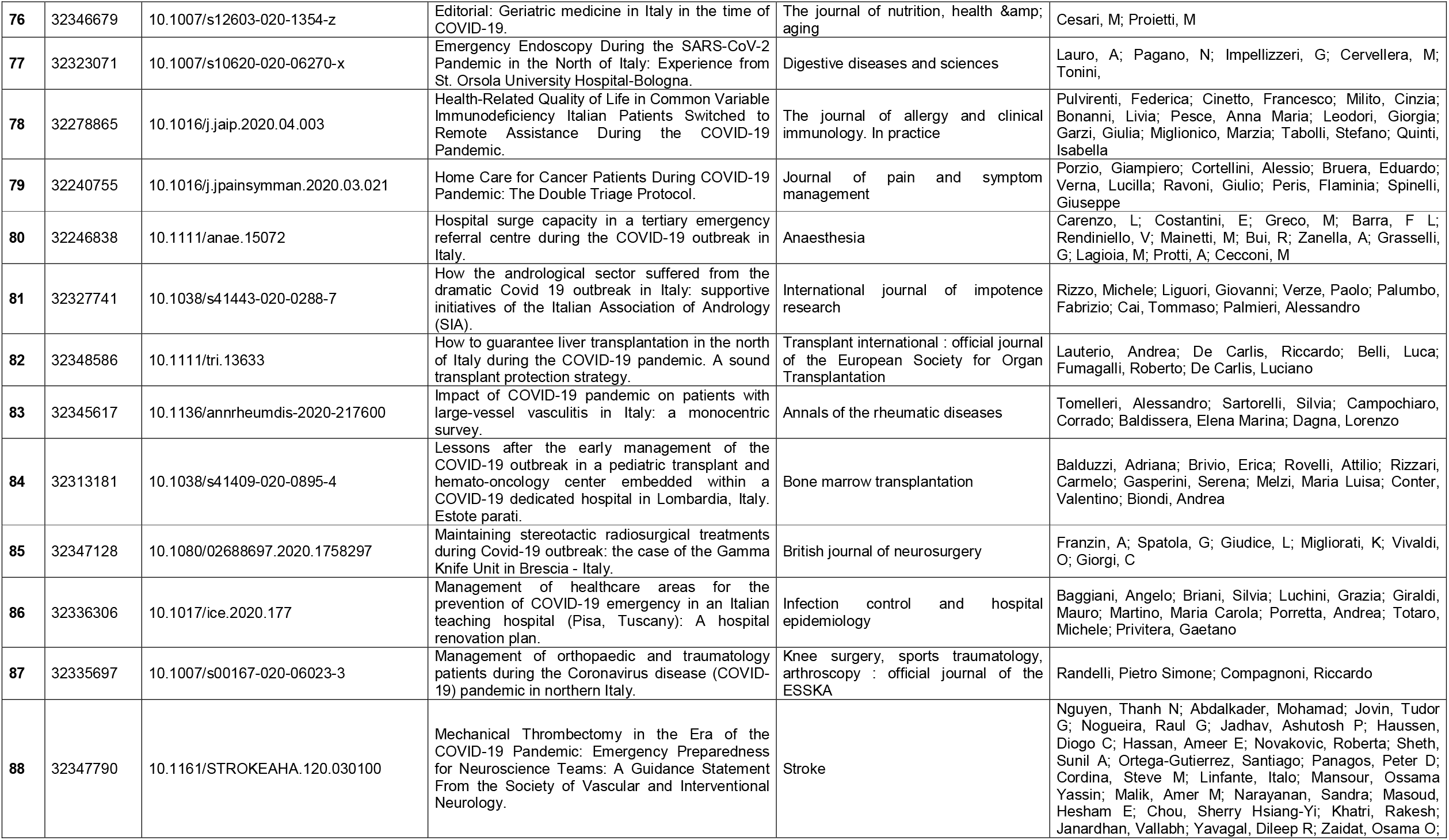
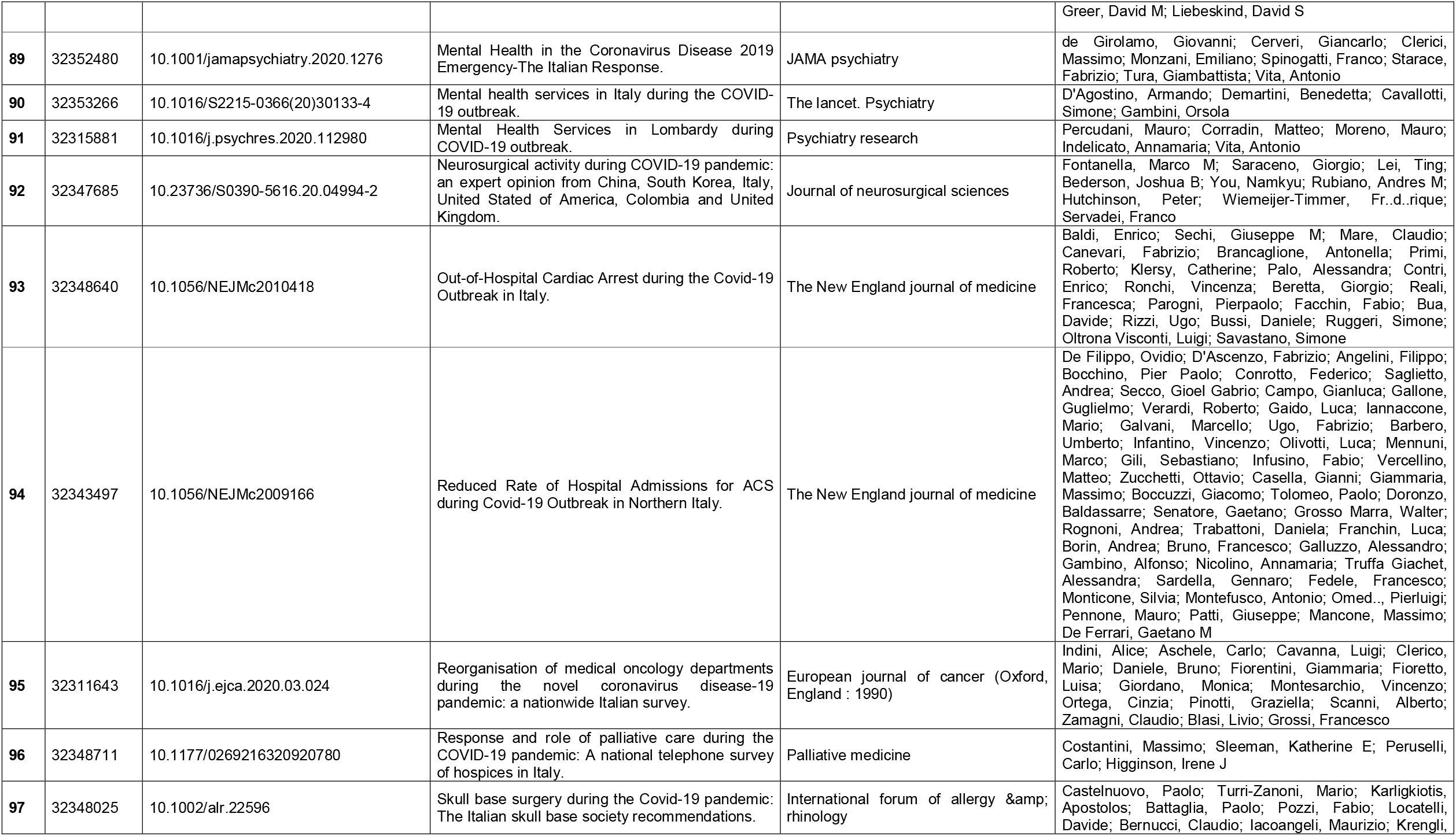
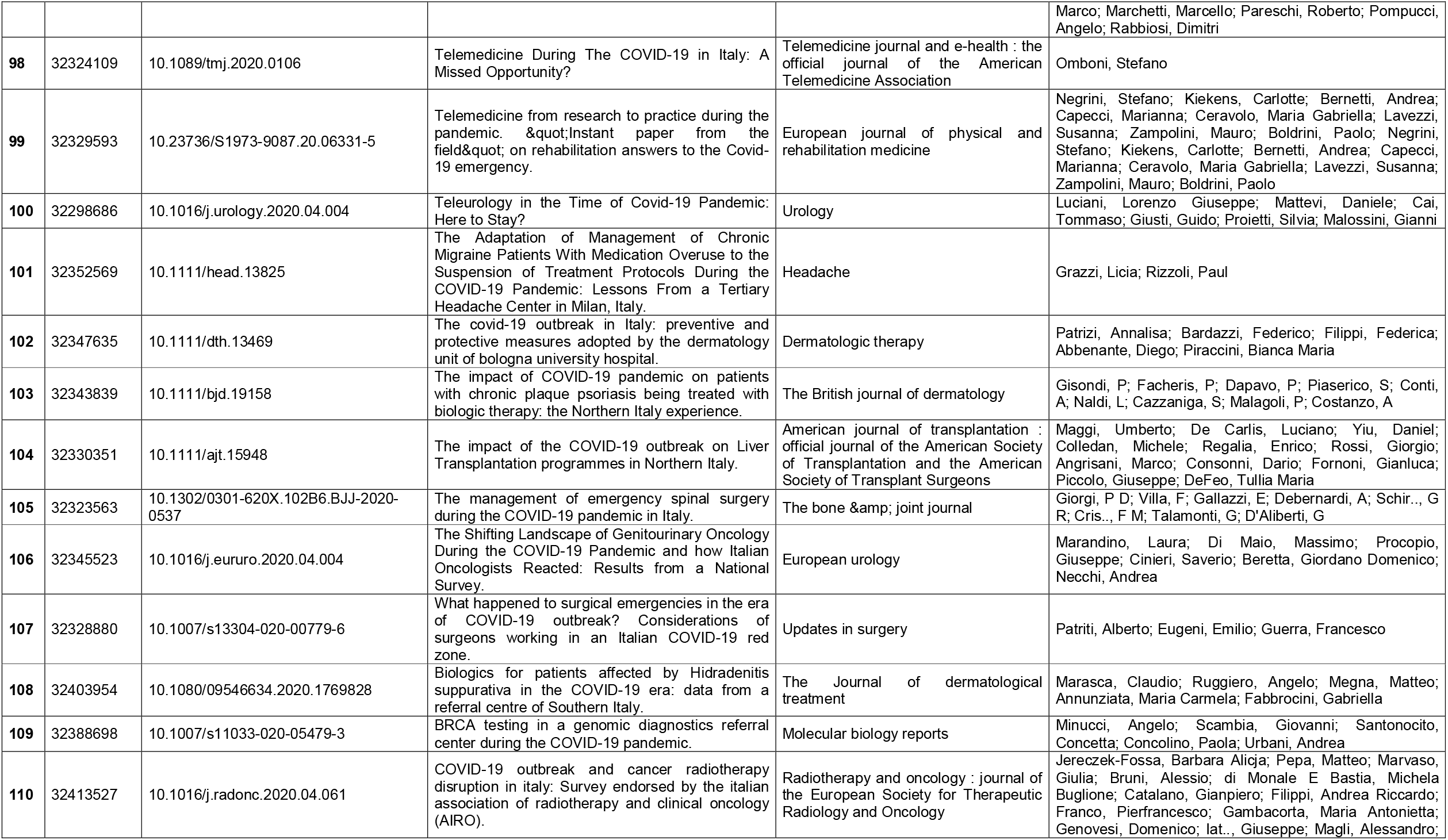
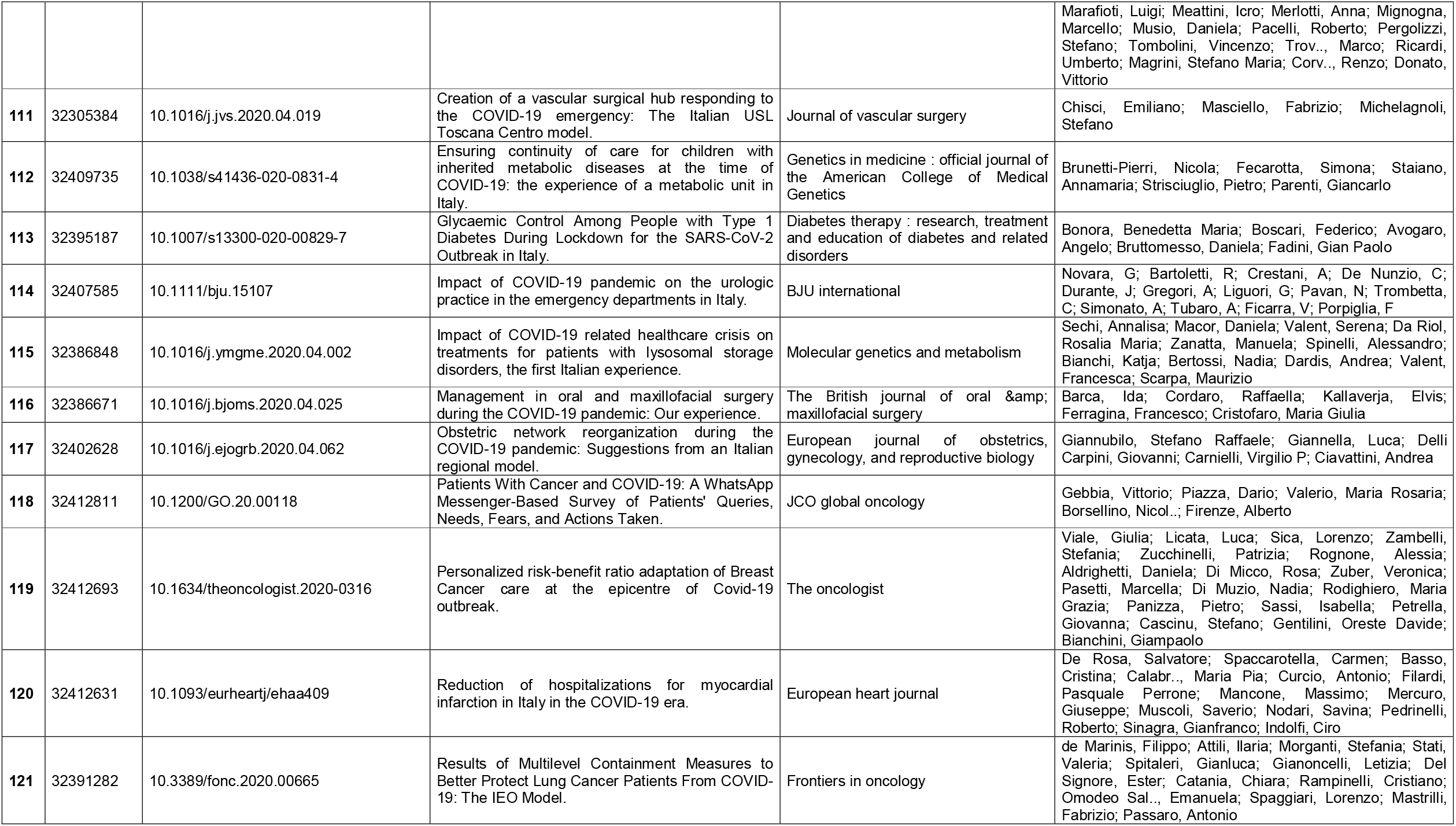
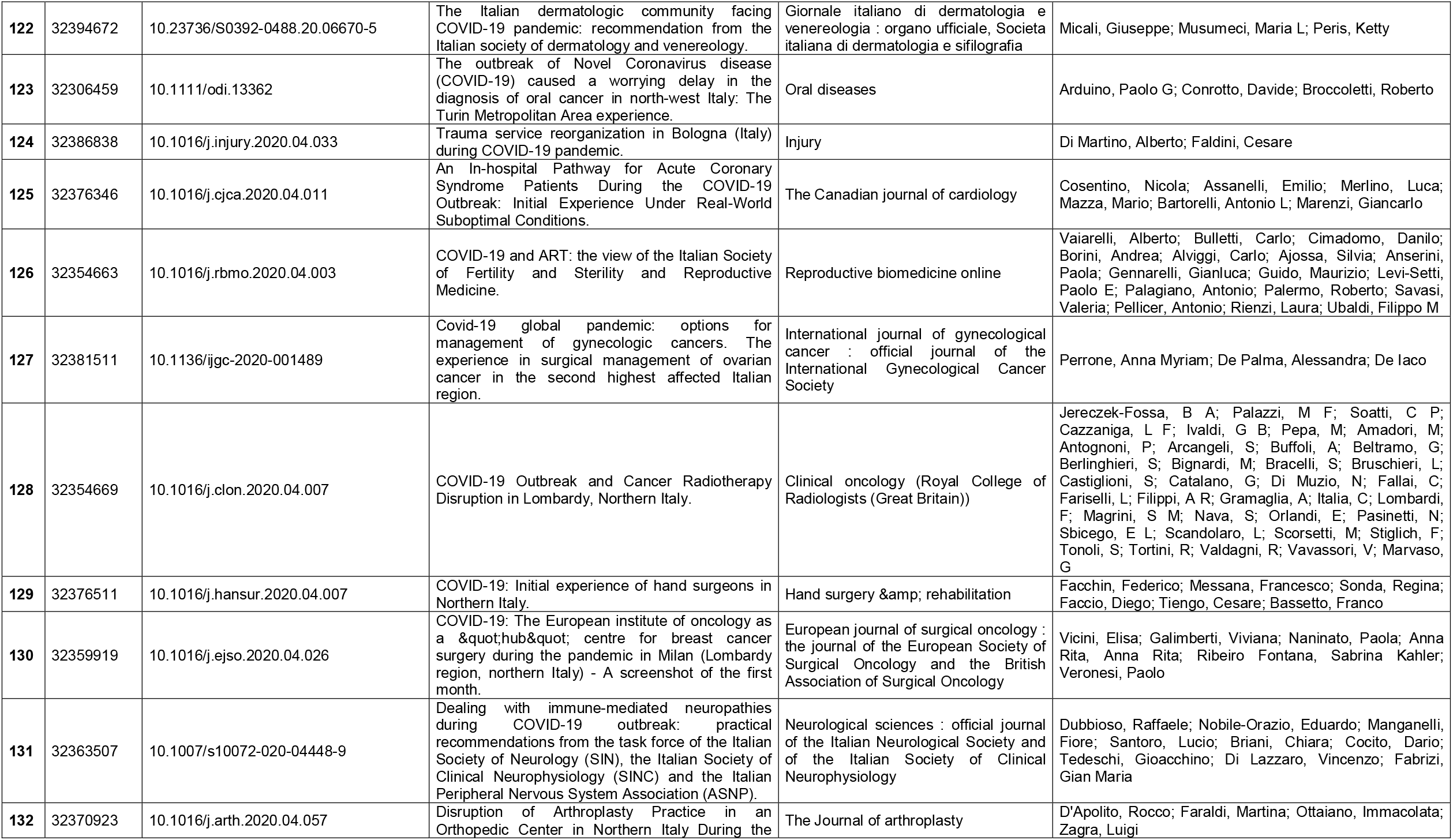
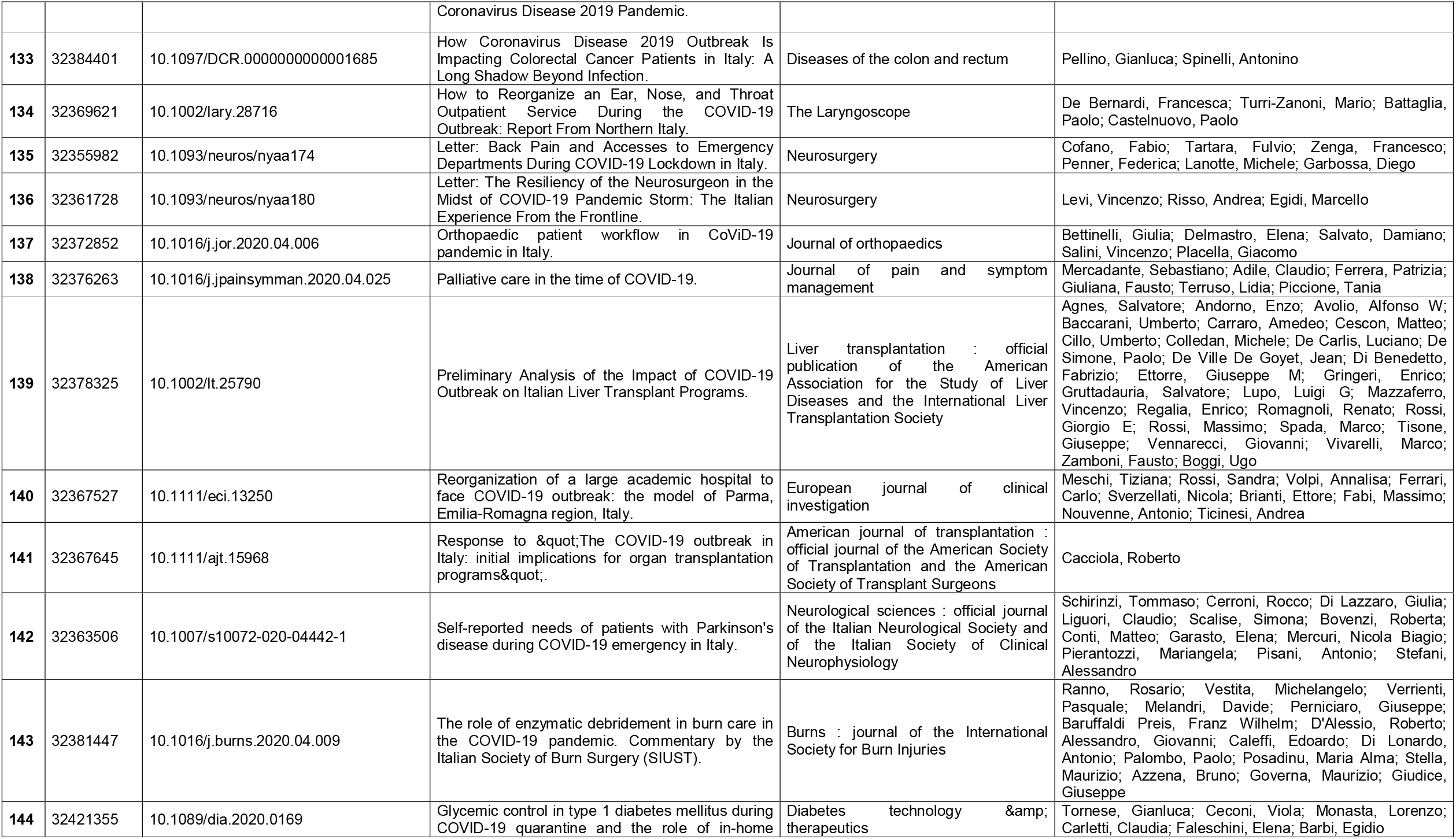
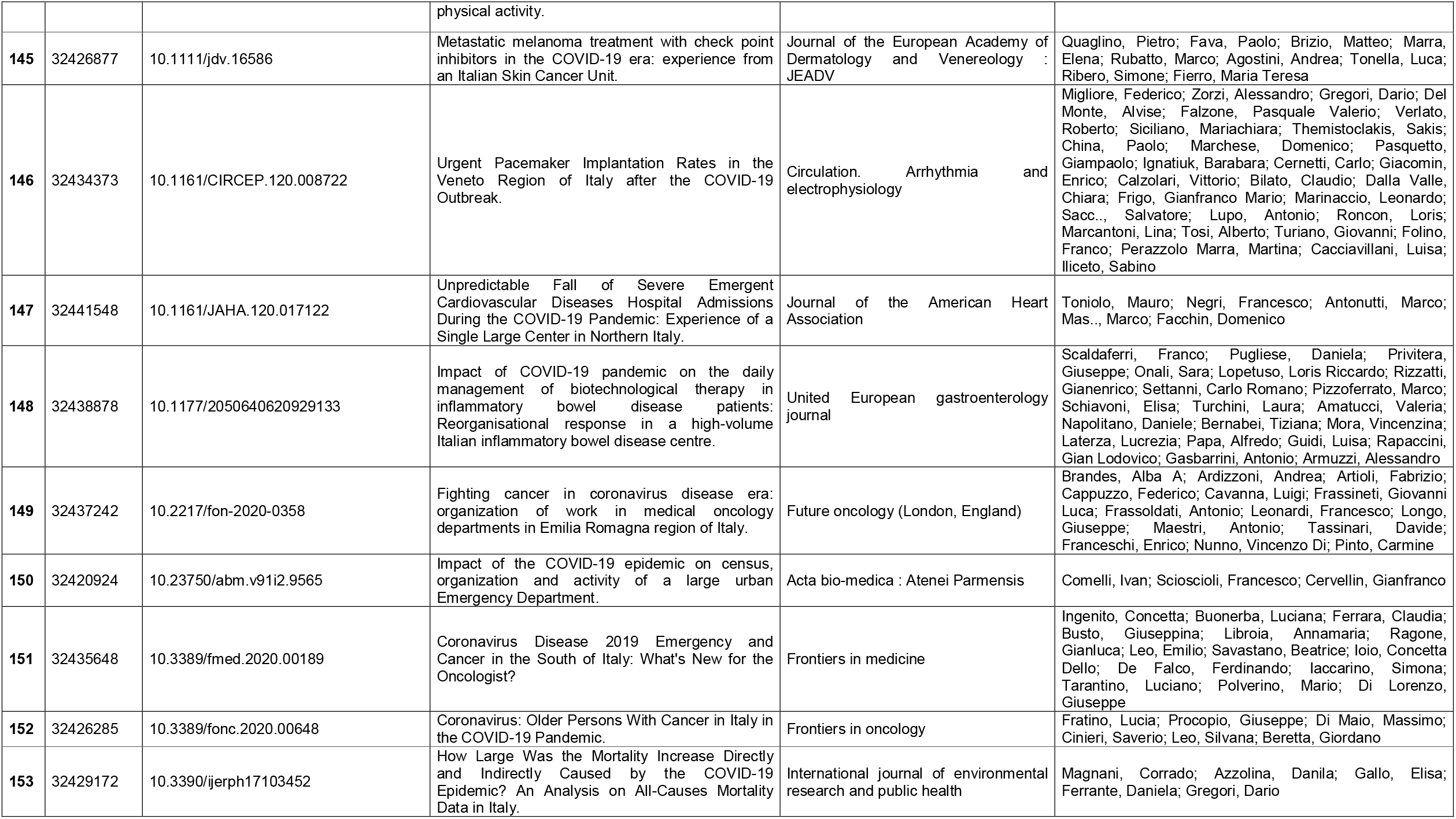
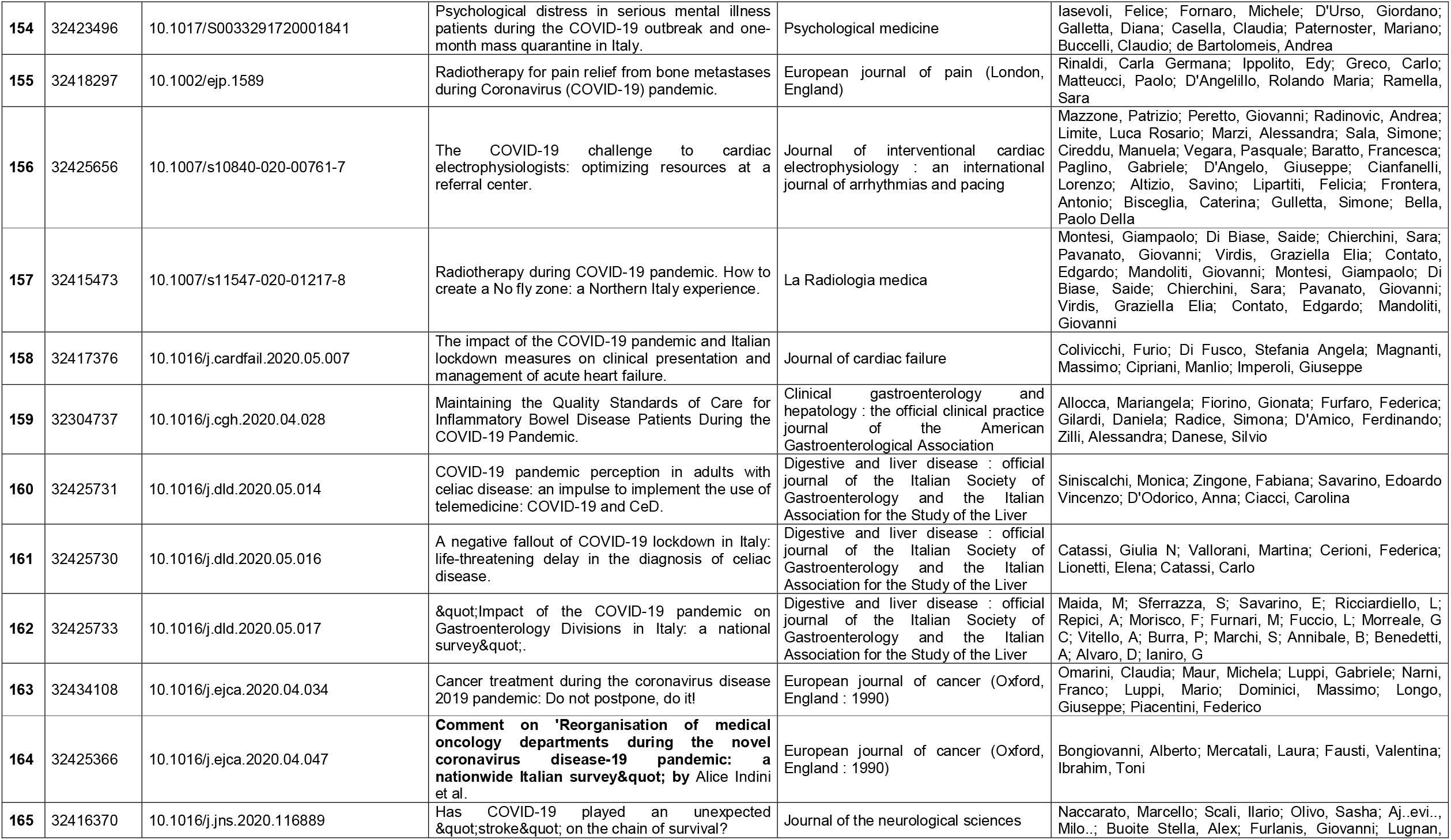
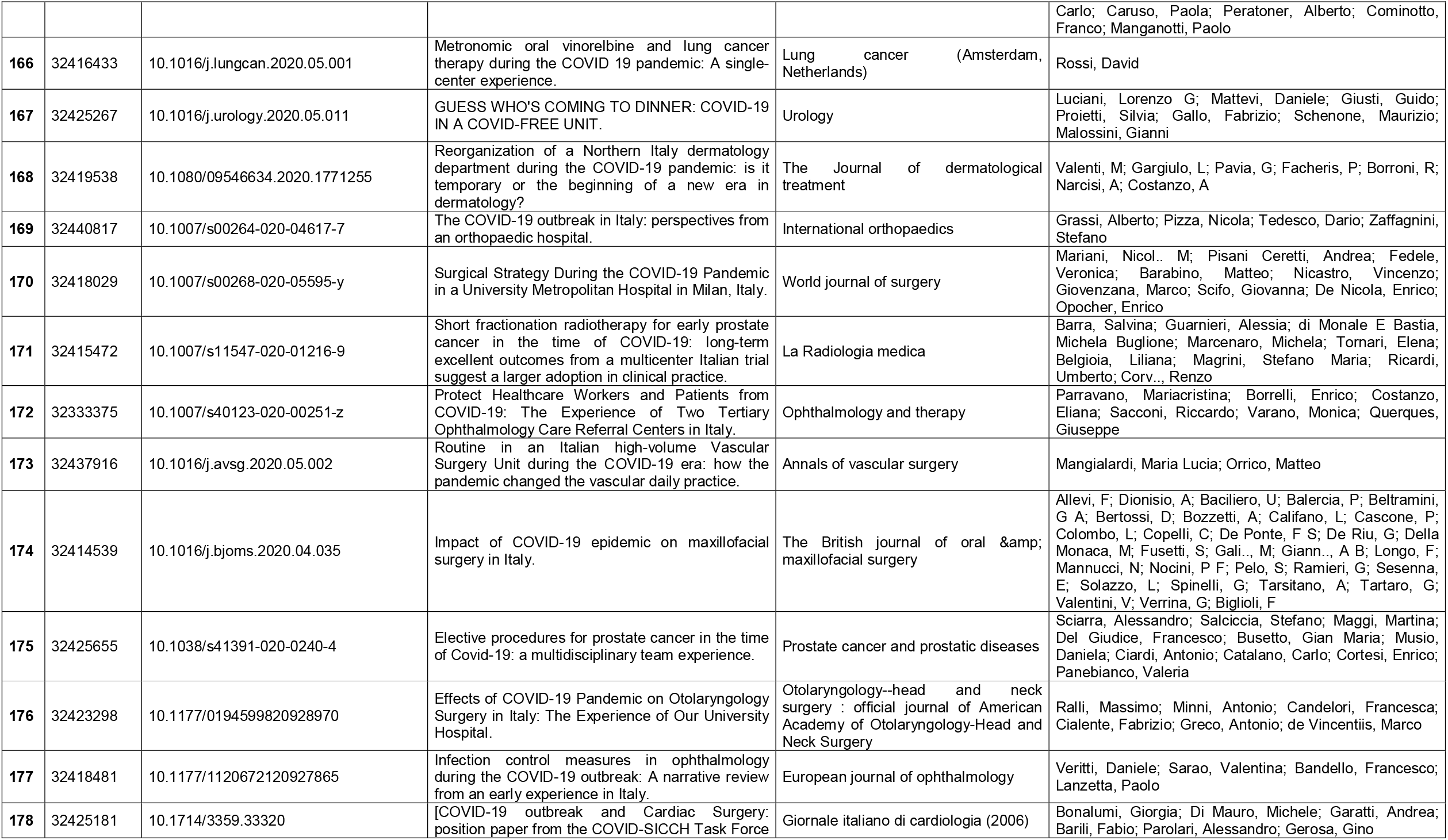
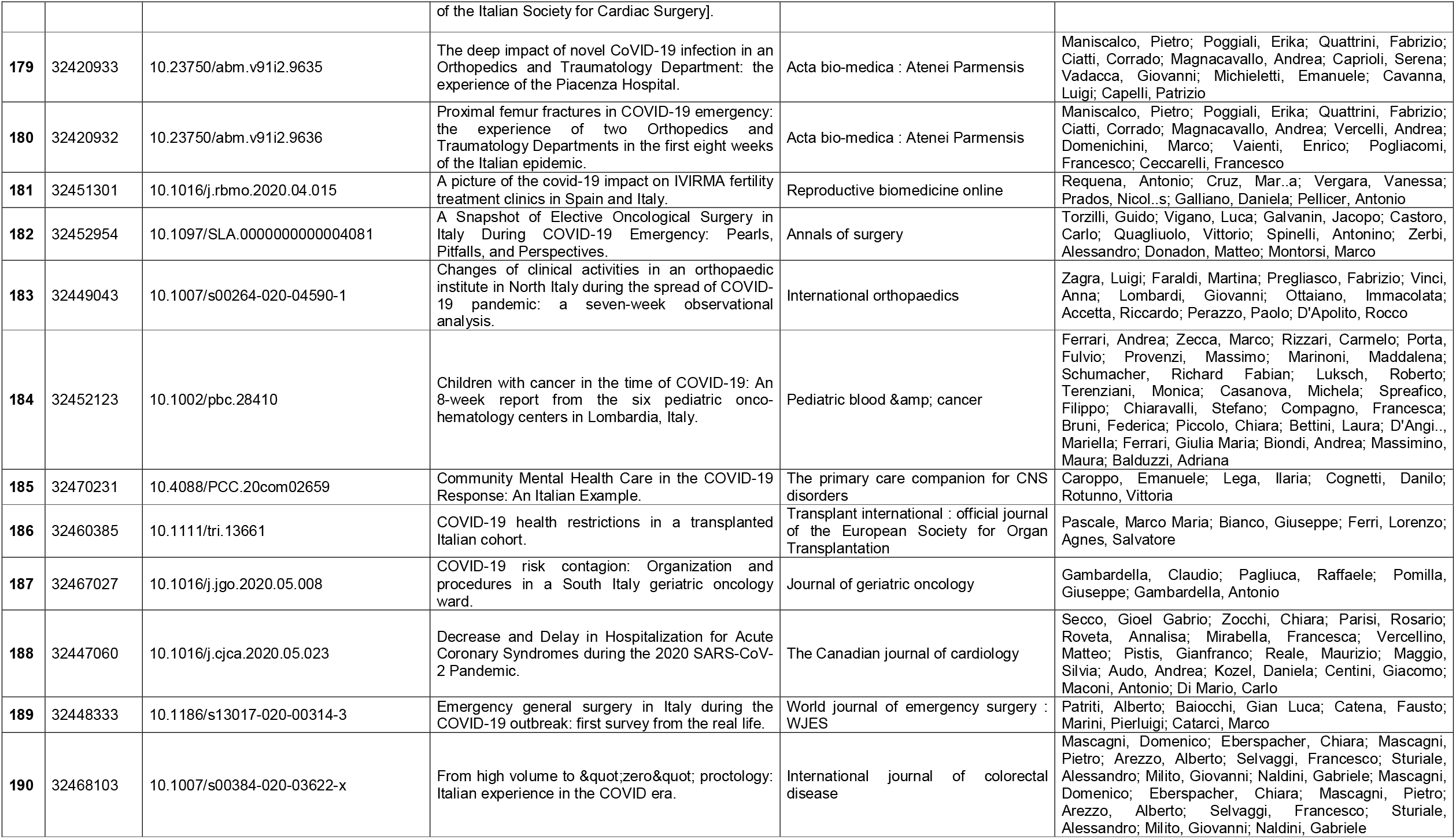
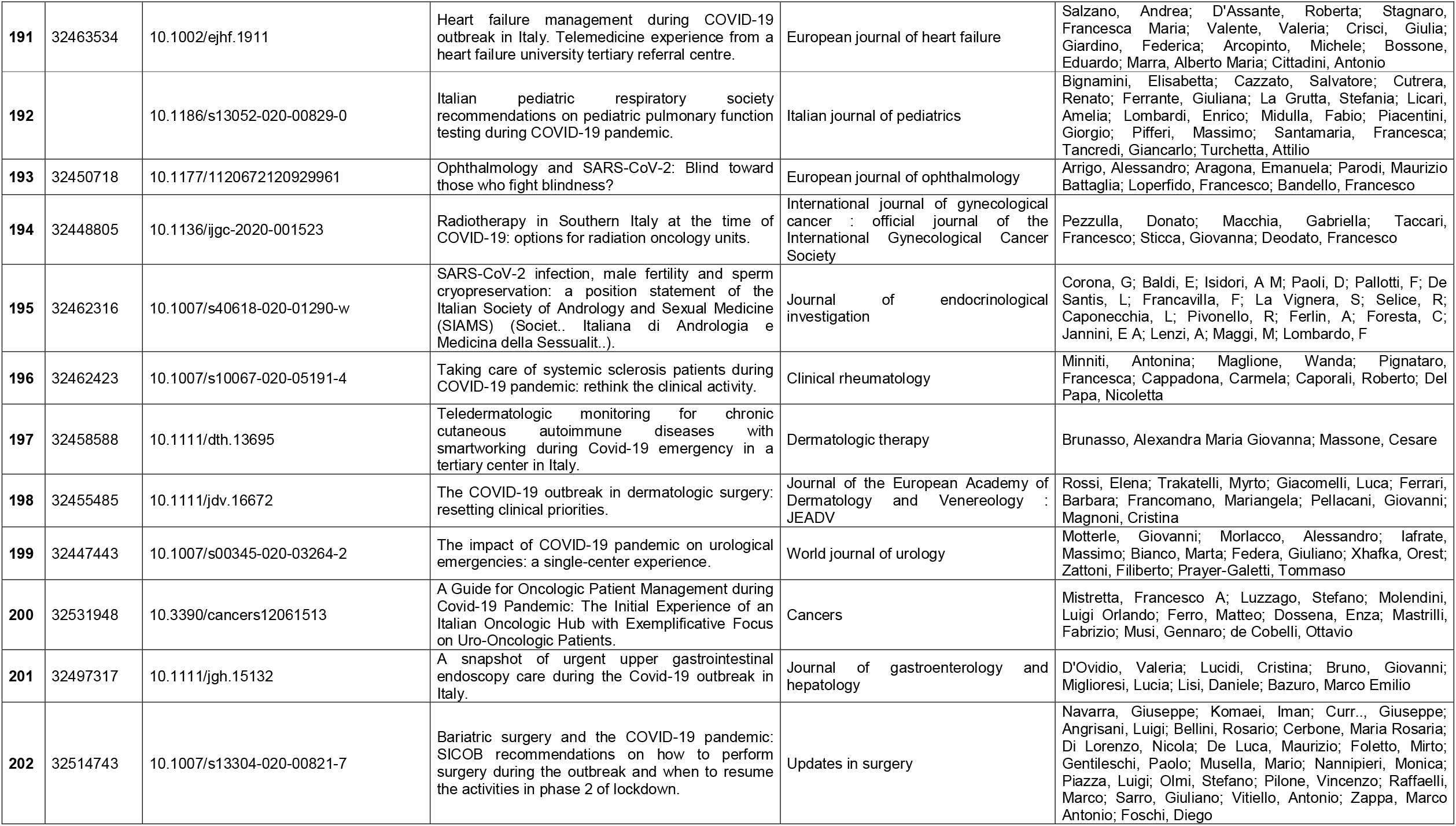
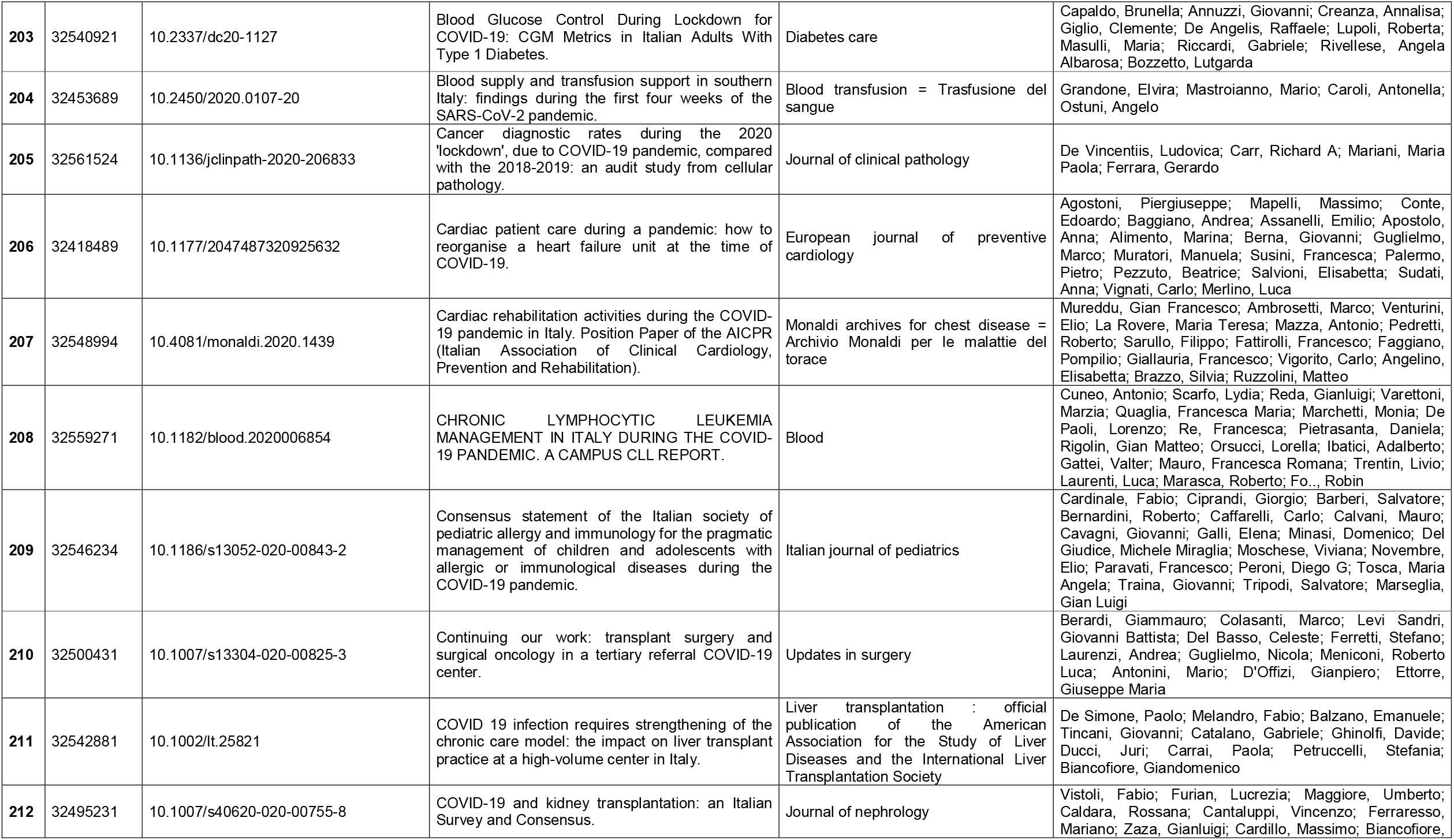
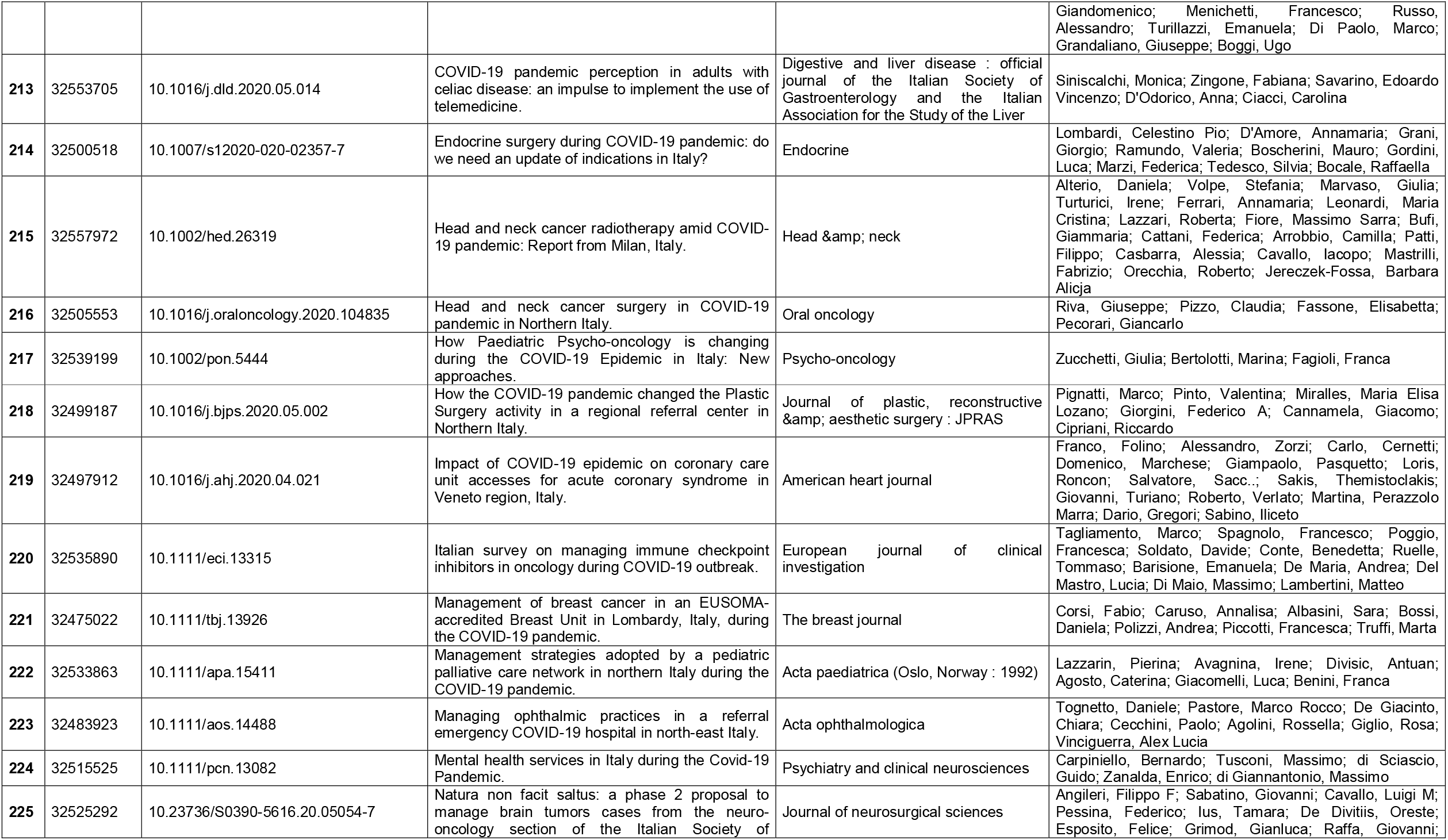
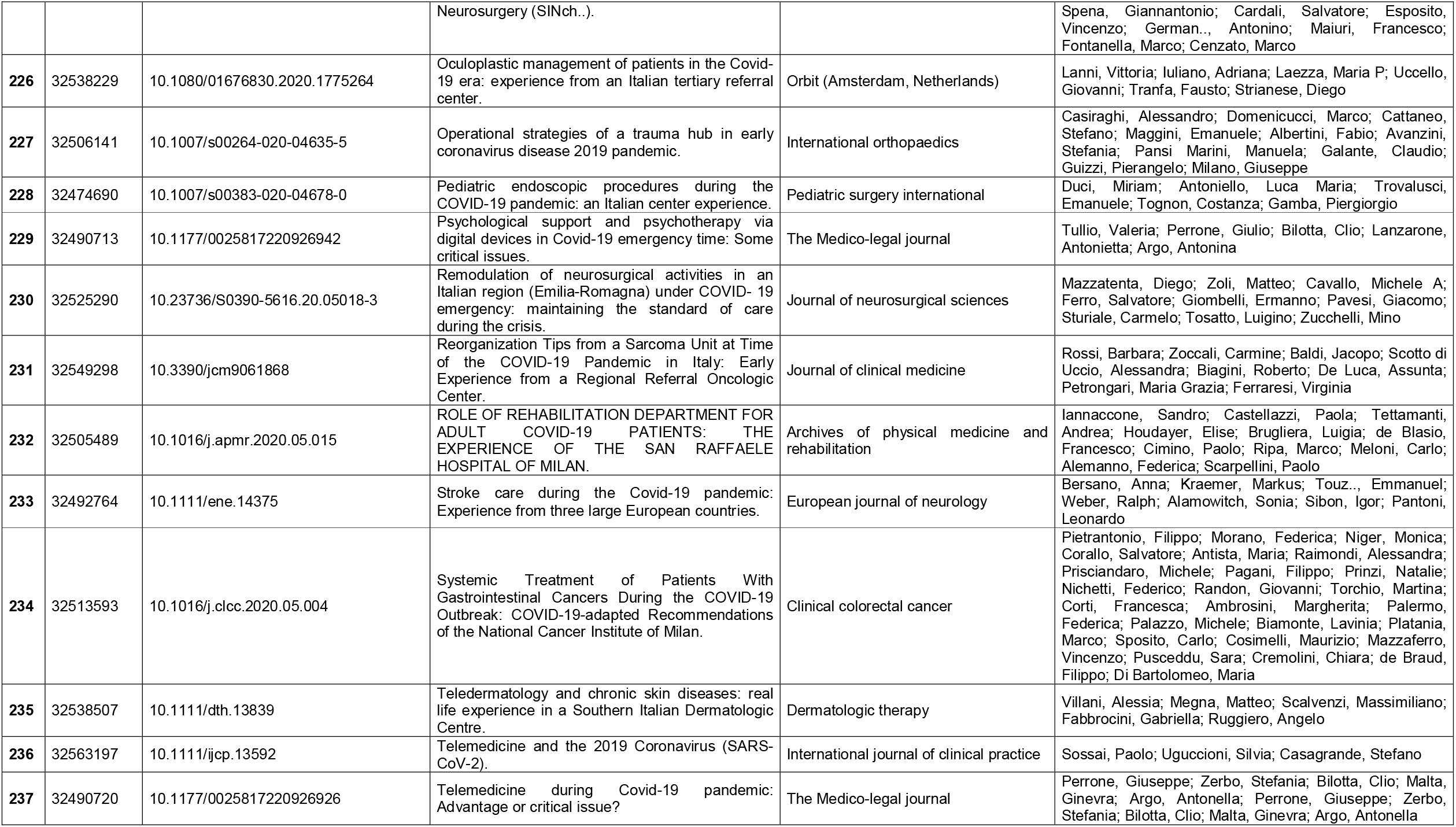
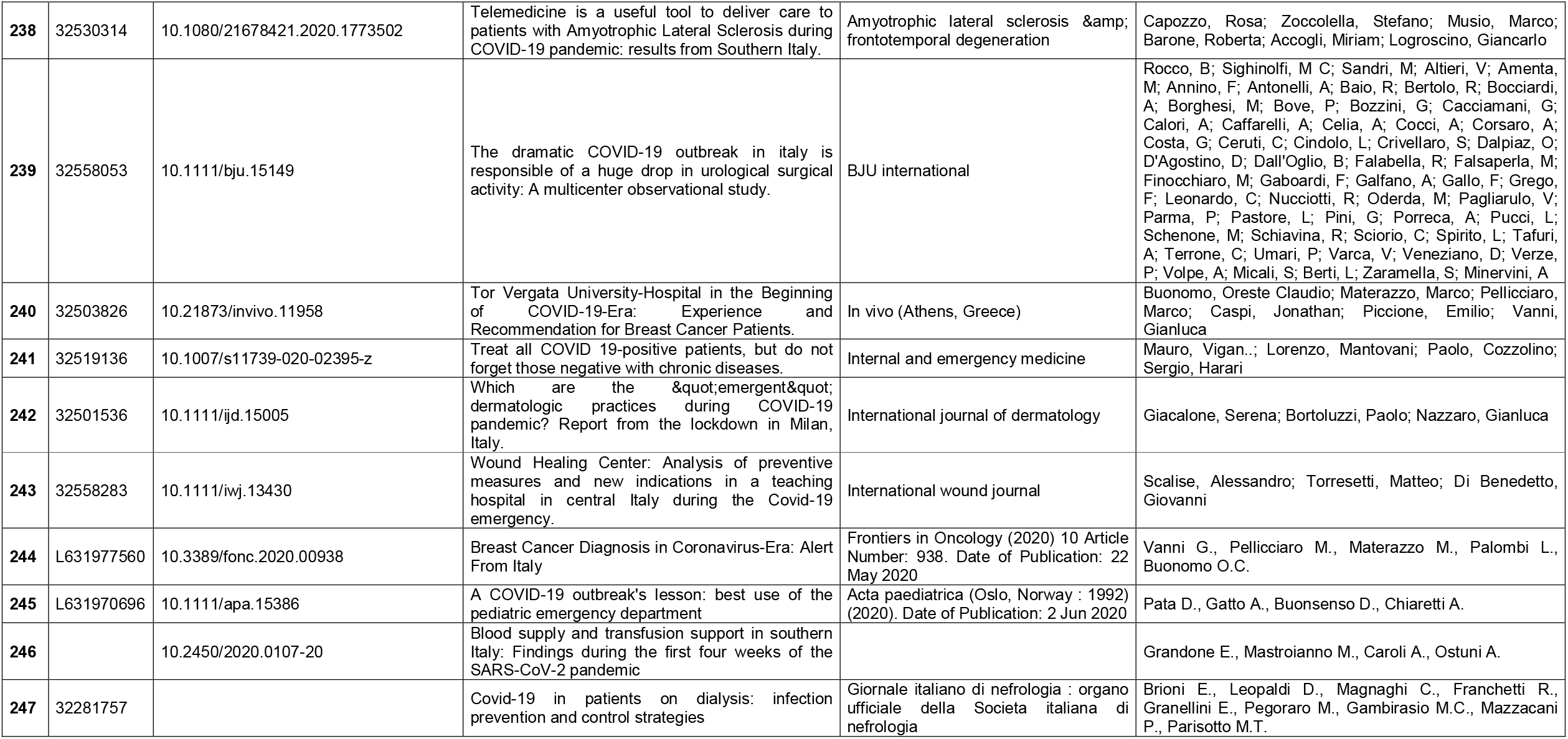
Studies included.

